# Effect of initiating an ARB- versus ACEI-based regimen on dementia risk, a target trial emulation of 2.5 million US Veterans

**DOI:** 10.64898/2026.07.05.26357173

**Authors:** Yizhe Xu, Jianlin Shi, Ryan M. Andrews, Catherine G. Derington, Tom H. Greene, Daniel O. Scharfstein, Ransmond O. Berchie, Mark A. Supiano, Jeff Williamson, Nicholas M. Pajewski, Jeremy J. Pruzin, Jaejin An, Jordana B. Cohen, Adam P. Bress

**Affiliations:** Department of Internal Medicine, Spencer Fox-Eccles School of Medicine, Division of Epidemiology, University of Utah, Salt Lake City, UT; Department of Epidemiology, Mailman School of Public Health, Columbia University, New York, NY; Division of Cardiology, University of Colorado School of Medicine, Aurora, CO; Adult & Child Center for Outcomes Research & Dissemination, University of Colorado Anschutz Medical Campus, Aurora, CO; Intermountain Healthcare Department of Population Health Sciences, Divisions of Health System Innovation and Research and Biostatistics, Spencer Fox-Eccles School of Medicine, University of Utah, Salt Lake City, UT; Geriatrics Division, Spencer Fox Eccles School of Medicine, University of Utah, Salt Lake City, UT; Center on Aging, University of Utah, Salt Lake City, UT; Department of Internal Medicine, Geriatric and Gerontology and the Sticht Center for Healthy Aging and Alzheimer’s Prevention, Wake Forest University School of Medicine, Winston-Salem, NC; Department of Biostatistics and Data Science, Wake Forest University School of Medicine, Winston-Salem, NC; Banner Alzheimer’s Institute, Phoenix, AZ; Department of Research and Evaluation, Kaiser Permanente Southern California, Pasadena, CA; Department of Medicine, Renal-Electrolyte Department of Medicine, Renal-Electrolyte and Hypertension Division, Perelman School of Medicine at the University of Pennsylvania, Philadelphia, PA; Department of Biostatistics, Epidemiology, and Informatics, Perelman School of Medicine, University of Pennsylvania, Philadelphia, PA; George E. Wahlen Department of Veterans Affairs Medical Center, Salt Lake City, UT

**Keywords:** antihypertensive medication, dementia, natural language processing, target trial emulation, Veteran

## Abstract

**Background:** Hypertension is a modifiable risk factor for dementia, yet the comparative effectiveness of angiotensin receptor blockers (ARBs) versus angiotensin converting enzyme inhibitors (ACEIs) on dementia risk remains uncertain.

**Objective:** To compare the risk of dementia and dementia-free death of ARB versus ACEI initiation among US Veterans with incident hypertension.

**Methods:** We conducted a retrospective target trial emulation using a new-user, active-comparator design among Veterans with incident hypertension. We analyzed longitudinal electronic health records from 2,577,000 individuals who initiated ARBs or ACEIs between 1/1/2000-12/31/2017, with up to five years of follow-up. The exposure was initiation of an ARB-based versus ACEI-based antihypertensive regimen. Co-primary outcomes were dementia, identified using natural language processing of clinical notes, and dementia-free death. We used inverse probability of treatment weights based on 66 pretreatment covariates to estimate the cumulative incidence of the outcomes for each treatment group. Weighted risk ratios and absolute risk differences through five years were computed with bootstrapped 95% CIs. Secondary outcomes included all-cause death and a composite of dementia or death, evaluated using a weighted Kaplan-Meier approach.

**Results:** Among 2,577,000 Veterans (mean age, 63 years; 4.5% female; 65% White; 15% Black), 10% initiated ARBs and 90% initiated ACEIs. Over five years of follow up, 6% developed dementia, 12% died without dementia, and 13% died overall. ARB initiation yielded consistently lower risk of dementia (risk ratio, 0.88; 95% CI, 0.83-0.93 at 6 months to 0.92; 95% CI, 0.90-0.94 at 5 years) and dementia-free death (risk ratio, 0.90; 95% CI, 0.86-0.96 at 6 months to 1.00; 95% CI, 0.98-1.01 at 5 years) than ACEI initiation. Effects on secondary outcomes were similar to those for primary outcomes. Greater protective dementia effects were observed in older and male Veterans and non-statin users, with similar effects on dementia-free death.

**Discussion:** Among US Veterans with incident treated hypertension, initiation of ARB versus ACEI antihypertensive regimen conveyed a modestly lower risk of dementia. Given the high prevalence of hypertension, these modest effects may confer meaningful population-level benefits on brain health. Future research estimating per-protocol effects using a more generalizable population is needed to confirm our findings.

## INTRODUCTION

Hypertension is a recognized modifiable risk factor for Alzheimer’s disease (AD) and AD-related dementias and may be particularly relevant for vascular dementia.^1–3^ Evidence from randomized trials and observational studies show that lowering blood pressure with antihypertensive medication can modestly reduce risk of cognitive impairment and dementia among older adults.^4–7^

Two major antihypertensive medication classes, angiotensin II receptor blockers (ARBs) and angiotensin-converting enzyme inhibitors (ACEIs), target the renin–angiotensin system but differ in their mechanisms.^8^ ARBs selectively antagonize angiotensin type 1 (AT1) receptors, thereby increasing the activation of angiotensin type 2 (AT2) and type 4 (AT4) receptors, a process linked to improved cerebral blood flow, reduced oxidative stress, and decreased neuroinflammation, key mechanisms potentially protective against late-life cognitive decline.^9–14^ In contrast, ACEIs reduce angiotensin II production, diminishing activation of all angiotensin II receptors, including the neuroprotective AT2 and AT4 subtypes.^12,15^

Despite emerging evidence that ARBs may provide additional cognitive benefits beyond blood pressure control, ACEIs are prescribed as first-line therapy nearly ten times more often.^16^ Given the high prevalence of hypertension, even modest advantages of ARBs could have far-reaching public health benefits by substantially reducing dementia burden and associated costs.^17–19^ Several observational studies have suggested lower dementia risk with ARBs compared with ACEIs,^20–25^ but most did not address competing risks, lacked power to evaluate subgroup effects, and relied on diagnostic codes, which underdiagnose dementia cases and mild cognitive impairment^26,27^ and often reflect racial and ethnic disparities in the ascertainment of cognitive impairment.^28,29^

In this study, we addressed these limitations by emulating a target trial among 2.5 million Veterans with newly diagnosed hypertension initiating either ARBs or ACEIs. By leveraging longitudinal data, natural language processing (NLP) to identify dementia cases from unstructured clinical notes, and causal inference methods to mitigate confounding by indication and account for the competing risk of death, we estimated the effects of ARB versus ACEI initiation on dementia and dementia-free death. These findings may inform whether prioritizing ARBs in hypertension management could improve cognitive outcomes in routine care.

## METHODS

### Specification of the Target Trial

We emulated a hypothetical target trial enrolling US Veterans with newly diagnosed hypertension between January 1, 2000, to December 31, 2017. Eligible Veterans would be at least 18 years old with no prior hypertension diagnosis, no prior ARB or ACEI use, and no history of dementia. Participants would be randomly assigned to initiate an ACEI or ARB in an open-label design and followed from randomization until the first event of dementia, loss to follow-up, or 5-year follow-up. The primary outcomes would be incident dementia and dementia-free death.

### Emulation of the Target Trial

We used longitudinal electronic health records (EHRs) from the Veterans Health Administration (VHA) to emulate the target trial with an observational, new-user, active-comparator, retrospective cohort design.^30,31^ Veterans were eligible if they (1) received a new hypertension diagnosis between January 1, 2000, and December 31, 2017; (2) had ≥1 primary care visit in the year before their first antihypertensive prescription (index date); (3) had no prior ARB or ACEI use before January 1, 2000; and (4) had no history of dementia at or before the index date. We excluded individuals with data inconsistencies (e.g., incorrect birth dates, multiple death dates), reidentification risk (e.g., age >90 years), and those who initiated neither or both ARB and ACEI, died, or were ineligible at the index date. This study adhered to the Strengthening the Reporting of Observational Studies in Epidemiology (STROBE) guideline and the Transparent Reporting of Observational Studies Emulating a Target Trial (TARGET) Statement.^32^ The protocol of the hypothetical target trial and its emulation steps are shown in **Table 1** and variable definitions are described in **eTable 3**.

**Table 1.** Hypothetical Target Trial and Its Emulation Framework. Comparison of the design components between the hypothetical target trial and its observational emulation using Veterans Health Administration data. The emulated trial followed Veterans from treatment initiation until dementia, death, loss to follow-up, or 5 years of follow up.

| Component | Hypothetical target trial | Emulated target trial |
| --- | --- | --- |
| Aim | To compare the intention-to-treat effects of initiating ARBs versus ACEIs on the risks of dementia and dementia-free death | Same. |
| Eligibility | 1. Age 18 years or older<br>2. No history of ARB or ACEI use prior to baseline<br>3. No history of dementia | Same. |
| Treatment strategies | 1. Initiate ARB treatment according to a clinical protocol<br>2. Initiate ACEI treatment according to a clinical protocol | Same. |
| Treatment assignment | Subjects are randomly assigned and are aware of the strategy (i.e., open label) | Participants are assigned to treatment strategies based on study-ascertained use of ARB or ACEI (using VHA outpatient pharmacy dispense data).<br>Randomization is emulated via use of propensity score in baseline treatment model (see Methods). |
| Follow-up | Participants are followed from randomization until earliest occurrence of: Dementia, death, loss to follow up (i.e., drop-out), or 5 years of follow up | Participants are followed from date of treatment initiation until earliest occurrence of: Dementia, death, loss to follow-up (i.e., drop-out), or 5 years of follow up. |
| Outcome | Primary: Dementia identified based on cognitive assessments and adjudicated by a panel of neurologists, neuropsychologists, geriatricians who are experienced in diagnosing cognitive syndromes | Primary: Dementia identified using natural language processing algorithms<br>Secondary: Composite of dementia and death; all-cause mortality; dementia identified using diagnosis code; dementia identified using diagnosis code and anti-dementia medication use |
| Causal contrast | The intention-to-treat effect of initiating ARBs versus ACEIs on 5-year dementia risk | Observational analog. |
| Statistical analysis | Intention-to-treat analysis: Comparison of the cumulative incidence at 5 years between treatment groups | Same, incorporating weights that are the estimated probabilities of baseline treatment assignment. |

### Standard Protocol Approvals, Registrations, and Patient Consents

The study was approved by the University of Utah review board, the Department of Veterans Affairs Institutional Review Board, and the VA Research & Development Committee. Because the analysis used existing EHR data from the Veterans Health Administration, a waiver of informed consent and a waiver of Health Insurance Portability and Accountability Act (HIPAA) authorization were granted in accordance with federal regulations.

### Treatment Strategies and Medication Exposure Assessment

The treatment strategies compared were initiation of an ARB-based versus ACEI-based antihypertensive regimen, as recorded in VHA outpatient pharmacy dispensing data. The index date was defined as the earliest ARB or ACEI prescription filling date between January 1, 2000, and December 31, 2017. Patients were categorized into the ARB or ACEI treatment strategies irrespective of use of other antihypertensive classes. Follow-up began on the index date and continued until dementia onset, death, loss to follow-up, or 5-year follow up, whichever came first.

### Outcome Identification and Ascertainment

We developed a hybrid NLP algorithm to identify dementia events from unstructured clinical notes. This approach integrated rule-based filtering, a binary support vector machine classifier, and a fine-tuned Bidirectional Encoder Representations from Transformers model to aggregate sentence-level predictions into patient-level dementia diagnoses. The algorithm achieved 84% sensitivity and 90% positive predictive value in identifying dementia cases^33^ and identified 2.7-fold more dementia cases than International Classification of Diseases (ICD) code-based ascertainment (378,833 versus 138,796) (**eMethods**). As secondary outcomes, we identified dementia events using ICD Ninth and Tenth Revision codes. Specifically, patients were classified as having dementia if they had ≥2 outpatient claims (≥7 days apart) or ≥1 inpatient claim for a dementia code in any position.

### Competing Risks

Because death can preclude observation of dementia, we specified dementia and dementia-free death as co-primary outcomes. For dementia, death was treated as the competing event; for dementia-free death, dementia was treated as the competing event, and deaths after dementia onset were not counted.

### Baseline Covariates

We preselected 66 baseline covariates with the potential to confound associations between ARB versus ACEI initiation and dementia or dementia-free death; covariates were extracted using published algorithms applied to the pre-index period (all covariates listed in **eMethods** and defined in **eTable 1**).

### Statistical Analysis

To address baseline covariate imbalance between treatment groups, we applied inverse probability (IP) of treatment weighting based on propensity scores (PSs) estimated from a logistic regression model that included baseline covariates and their two-way interactions with seven prespecified subgroup variables.^34–36^ To maintain model parsimony, we applied the least absolute shrinkage and selection operator to select important covariate-subgroup interactions (**eMethods**).^37^ Extreme weights were truncated at the 99^th^ percentile. We assessed covariate balance before and after weighting using absolute standardized mean differences (ASMD), with ASMD<0.1 indicating acceptable balance.

To estimate intention-to-treat (ITT) effects of ARB versus ACEI initiation on dementia and dementia-free death, we applied a competing risk framework.^38–40^ For each treatment group, we estimated the cumulative incidence of dementia at time t using an IP weighted Aalen-Johansen estimator (**eMethods**). We computed risk ratios (RRs) and risk differences (RDs) through 5 years, assuming noninformative loss to follow-up.

To address missing baseline covariate data, we conducted multiple imputation by chained equations, generating 10 imputed data sets.^41^ For laboratory variables unlikely to be missing at random based on experts’ domain knowledge, a separate “unknown” category was created.

For each completed dataset, we estimate standard errors of appropriately transformed effect estimates using nonparametric bootstrap (200 re-samples). We used Rubin’s combining rules^42^ to pool these estimates and compute standard errors. We derived 95% confidence intervals (CIs) assuming normality and back-transformed them to effect scales of interest.

### Secondary, Subgroup, and Sensitivity Analyses

We used the same IP weights above to estimate cause-specific hazard ratios (HR) by year. These cause-specific HRs complement the RRs by factoring dementia event counts by the number of surviving patients who remain at risk for dementia within each treatment group over follow-up. To assess the robustness of our dementia identification, we repeated the primary analysis for secondary dementia outcomes identified by ICD codes alone and by ICD codes plus anti-dementia medication use. In addition, we estimated RRs and RDs for the secondary composite outcome of dementia or death and for all-cause death using an IP-weighted Kaplan-Meier estimator.

We also evaluated effect modification by repeating the primary analysis across clinically relevant subgroups: age (<60 versus ≥60 years), sex (female versus male), race (Black versus non-Black), systolic blood pressure (SBP) (<140 versus ≥140 mm Hg), diabetes status, number of antihypertensive agents (<2 versus ≥2), and statin use. We used the estimated PSs from the primary analysis and performed Wald tests to assess whether treatment effects differ across patient subgroups.

For sensitivity analyses, we repeated all analyses using overlap weighting, which places larger weights on patients with PSs near 0.5.^43^ In our setting where ARBs were used roughly one-tenth as often as ACEIs, the average treatment effect in the overlap population approximates the “average treatment effect in the treated” (ATT). (**eMethods**).

All analyses were conducted using the Veterans Informatics and Computing Infrastructure (VINCI) in SAS Grid Enterprise 8.0x (SAS Institute) and R version 4.4.0 (R Foundation for Statistical Computing).

### Data Availability

The EHR data used in this study are available in the Veterans Health Administration.

## RESULTS

### Veteran Characteristics

Of 2,577,000 Veterans, 247,802 (10%) initiated an ARB and 2,329,198 (90%) initiated an ACEI. Continuous (no gap in fills greater than 9 months) 5-year use occurred in 46% of ARB initiators and 42% of ACEI initiators (**eFigure 12**). Before weighting, the mean (SD) age was 63 (12) years, 5% were female, 5% were Hispanic, 15% Black, and 65% White (**eFigure 1**). Compared with ACEI initiators, ARB initiators entered the cohort later (median index year 2010 versus 2007), were older, had higher household income, more cardiovascular disease, more heart failure, and fewer current smokers (**Table 2 and eTable 4**). After IP weighting, all baseline covariates were well balanced (ASMD<0.10) (**eFigure 3**), with broad overlap in treatment propensity scores (**eFigure 2**). **eTable 5** provides additional pre- and post-overlap weighting characteristics, and **eTable 15** details missing data.

**Table 2.** Baseline Characteristics of Veterans Before and After Inverse Probability Weighting. Continuous variables are summarized as mean (SD) or median (IQR), and categorical variables as counts (percentages). Absolute standardized mean differences (ASMD) <0.10 indicate acceptable balance. Full characteristics are provided in eTable 4.

| Variable | Before IP Weighting |  |  | After IP Weighting |  |  |
| --- | --- | --- | --- | --- | --- | --- |
|  | ARB<br>N = 247,802 | ACEI<br>N = 2,329,198 | ASMD | ARB | ACEI | ASMD |
| **Age (years)** | 65 (12) | 63 (12) | 0.20 | 64 (12) | 63 (12) | 0.08 |
| **Sex** |  |  | 0.05 |  |  | 0.01 |
| Female | 13,616 (5.5%) | 101,354 (4.4%) |  | 10,394 (4.6%) | 103,066 (4.4%) |  |
| Male | 234,186 (95%) | 2,227,844 (96%) |  | 215,530 (95%) | 2,213,566 (96%) |  |
| **Race/Ethnicity** |  |  | 0.06 |  |  | 0.048 |
| Hispanic | 8,973 (3.6%) | 107,160 (4.6%) |  | 8,979 (4%) | 104,558 (4.5%) |  |
| Non-Hispanic Black | 35,398 (14%) | 352,508 (15%) |  | 32,095 (14%) | 348,627 (15%) |  |
| Non-Hispanic White | 162,101 (65%) | 1,501,069 (64%) |  | 146,199 (65%) | 1,494,293 (65%) |  |
| Other | 41,330 (17%) | 368,461 (16%) |  | 38,650 (17%) | 369,155 (16%) |  |
| **Body mass index, kg/m<sup>2</sup>** | 31.2 (5.9) | 30.5 (6.0) | 0.12 | 30.7 (5.8) | 30.5 (6) | 0.03 |
| **Systolic blood pressure, mm Hg** | 141 (18) | 144 (18) | 0.14 | 143 (18) | 144 (18) | 0.02 |
| **Cardiovascular disease** | 62,336 (25%) | 245,541 (11%) | 0.39 | 26,991 (12%) | 265,521 (11%) | 0.01 |
| **Chronic kidney disease** | 9,743 (3.9%) | 65,539 (2.8%) | 0.06 | 8,544 (3.8%) | 68,641 (3%) | 0.05 |
| **Frailty<sup>1</sup>** |  |  | 0.03 |  |  | 0.04 |
| Frail | 7,528 (3.0%) | 60,055 (2.6%) |  | 7,491 (3.3%) | 62,650 (2.7%) |  |
|  | ARB<br>N = 247,802 | ACEI<br>N = 2,329,198 | ASMD | ARB | ACEI | ASMD |
| Non-frail | 240,274 (97%) | 2,269,143 (97%) |  | 218,432 (97%) | 2,253,982 (97%) |  |

<sup>1</sup>31-item index used to define frailty. Ratio of the sum of the number of health deficits to the total number of health factors evaluated

### Primary Outcome

Nearly all Veterans (>99.9%) were followed for at least 5 years. Among ARB initiators, 13,550 developed dementia (12.2/1000 person-years [PY]) and 30,404 died dementia-free (27.4/1000 PY), compared with 138,636 dementia cases (13.3/1000 PY) and 275,015 dementia-free deaths (26.4/1000 PY) among ACEI initiators. IP-weighted cumulative incidence of dementia was consistently lower under ARB versus ACEI initiation, with RRs attenuating from 0.88 (95% CI, 0.83-0.93) at 6 months to 0.92 (95% CI, 0.90-0.94) at 5 years and RDs increasing modestly from −0.001 (95% CI, −0.001 to −0.001) at 6 months to −0.005 (95% CI, −0.006 to −0.003) at 5 years. The number needed to treat to prevent one dementia event over 5 years was 200. Dementia-free death also initially favored ARB on the RR scale but fully attenuated by 5 years, with negligible RDs (**Figure 1** and **Table 3**). In secondary analysis, IP-weighted cause-specific HRs for dementia similarly attenuated over time (**eTable 6**).

**Figure 1.**
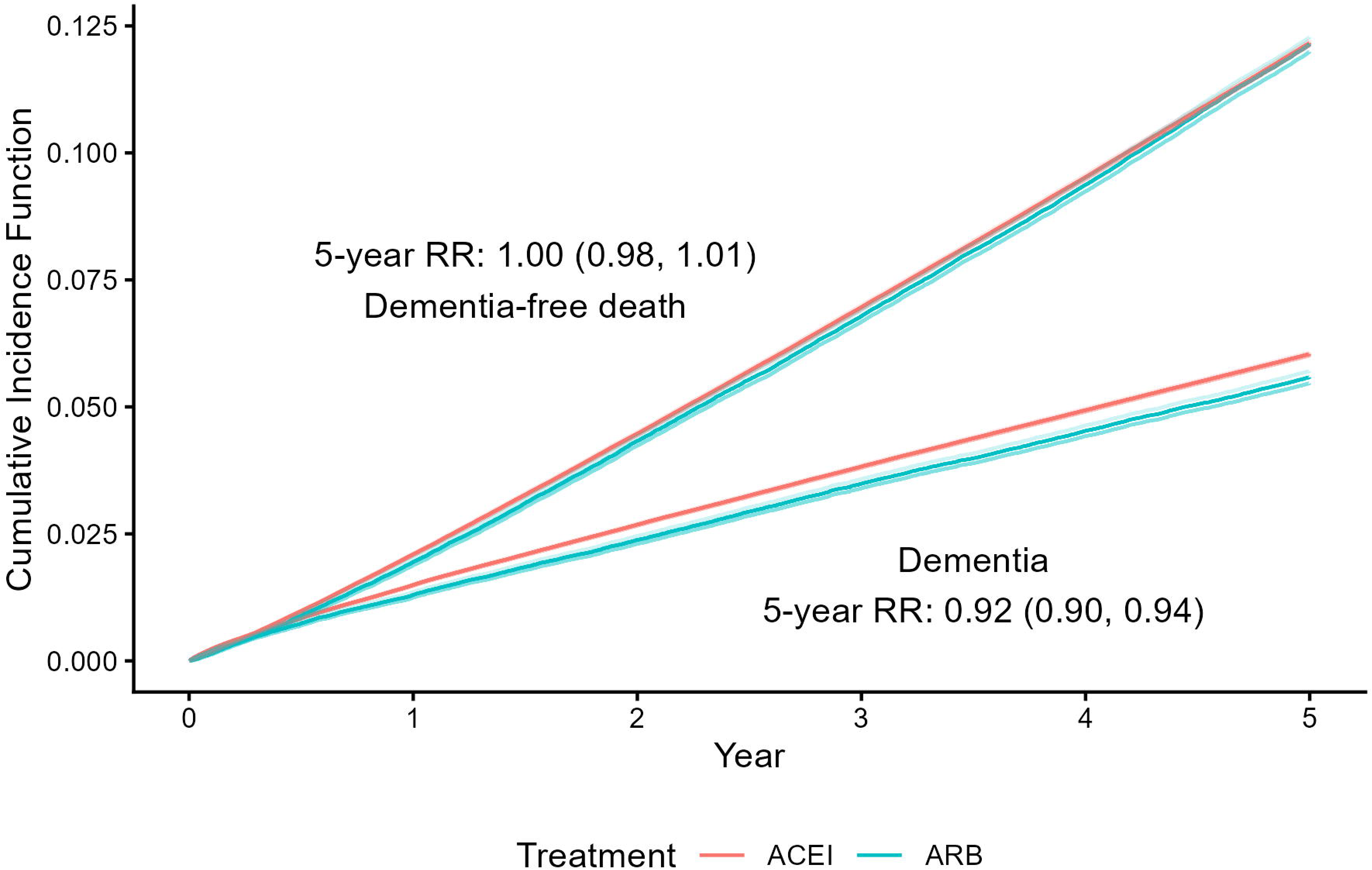
Inverse Probability Weighted Cumulative Incidence of Dementia and Dementia-free Death. Cumulative incidence curves show the intention-to-treat probabilities of dementia and dementia-free death among ARB and ACEI initiators, estimated using a competing risk framework. Dementia was identified using a validated NLP algorithm. Shaded areas indicate 95% confidence intervals.

**Table 3.** Inverse Probability Weighted Risks for Primary and Secondary Outcomes among ARB versus ACEI Initiators. Absolute risks, risk ratios, and risk differences for dementia, dementia-free death, the composite of dementia or death, and all-cause mortality at 5 years. Dementia was identified using a validated natural language processing (NLP) algorithm.

| Outcome | Absolute risk (95% CI) |  | Risk Ratio<br>(95% CI) | Risk Difference<br>(95% CI) |
| --- | --- | --- | --- | --- |
|  | ARB initiators<br>(n = 225,923) | ACEI initiators<sup>1</sup><br>(n = 2,316,632) |  |  |
| Primary Outcome |  |  |  |  |
| Dementia |  |  |  |  |
| 0.5-year | 0.007 (0.007, 0.008) | 0.008 (0.008, 0.009) | 0.879 (0.830, 0.931) | -0.001 (-0.001, -0.001) |
| 1-year | 0.013 (0.012, 0.014) | 0.015 (0.015, 0.015) | 0.870 (0.833, 0.909) | -0.002 (-0.003, -0.001) |
| 2-year | 0.024 (0.023, 0.025) | 0.027 (0.027, 0.027) | 0.887 (0.858, 0.918) | -0.003 (-0.004, -0.002) |
| 3-year | 0.035 (0.034, 0.036) | 0.038 (0.038, 0.039) | 0.912 (0.888, 0.937) | -0.003 (-0.004, -0.002) |
| 4-year | 0.045 (0.044, 0.046) | 0.049 (0.049, 0.050) | 0.918 (0.896, 0.940) | -0.004 (-0.005, -0.003) |
| 5-year | 0.056 (0.055, 0.057) | 0.060 (0.060, 0.061) | 0.924 (0.904, 0.944) | -0.005 (-0.006, -0.003) |
| Dementia-free death<sup>2</sup> |  |  |  |  |
| 0.5-year | 0.009 (0.009, 0.009) | 0.010 (0.010, 0.010) | 0.907 (0.860, 0.957) | -0.001 (-0.001, 0.000) |
| 1-year | 0.019 (0.019, 0.020) | 0.021 (0.021, 0.021) | 0.927 (0.895, 0.961) | -0.002 (-0.002, -0.001) |
| 2-year | 0.043 (0.042, 0.044) | 0.045 (0.044, 0.045) | 0.968 (0.946, 0.990) | -0.001 (-0.002, 0.000) |
| 3-year | 0.068 (0.067, 0.069) | 0.070 (0.069, 0.070) | 0.975 (0.958, 0.992) | -0.002 (-0.003, -0.001) |
| 4-year | 0.094 (0.092, 0.095) | 0.095 (0.095, 0.096) | 0.984 (0.970, 0.999) | -0.002 (-0.003, 0.000) |
| 5-year | 0.121 (0.120, 0.123) | 0.122 (0.121, 0.122) | 0.997 (0.984, 1.010) | 0.000 (-0.002, 0.001) |
| Secondary outcomes |  |  |  |  |
| Composite of dementia and death |  |  |  |  |
| 0.5-year | 0.016 (0.016, 0.017) | 0.018 (0.018, 0.019) | 0.894 (0.862, 0.928) | -0.002 (-0.003, -0.001) |
| 1-year | 0.033 (0.032, 0.033) | 0.036 (0.036, 0.036) | 0.904 (0.880, 0.928) | -0.003 (-0.004, -0.003) |
| 2-year | 0.067 (0.066, 0.068) | 0.072 (0.071, 0.072) | 0.938 (0.920, 0.956) | -0.004 (-0.006, -0.003) |
| 3-year | 0.103 (0.101, 0.104) | 0.108 (0.108, 0.108) | 0.953 (0.939, 0.967) | -0.005 (-0.007, -0.004) |
| 4-year | 0.139 (0.137, 0.141) | 0.145 (0.144, 0.145) | 0.961 (0.950, 0.973) | -0.006 (-0.007, -0.004) |
| 5-year | 0.177 (0.175, 0.179) | 0.182 (0.182, 0.183) | 0.973 (0.962, 0.984) | -0.005 (-0.007, -0.003) |
| All-cause death |  |  |  |  |
| 0.5-year | 0.009 (0.009, 0.010) | 0.010 (0.010, 0.010) | 0.897 (0.851, 0.945) | -0.001 (-0.002, -0.001) |
| 1-year | 0.020 (0.020, 0.021) | 0.022 (0.022, 0.022) | 0.920 (0.889, 0.953) | -0.002 (-0.002, -0.001) |
| 2-year | 0.046 (0.045, 0.047) | 0.048 (0.048, 0.049) | 0.958 (0.938, 0.979) | -0.002 (-0.003, -0.001) |
| 3-year | 0.074 (0.073, 0.075) | 0.077 (0.076, 0.077) | 0.965 (0.948, 0.981) | -0.003 (-0.004, -0.001) |
| 4-year | 0.104 (0.102, 0.105) | 0.107 (0.106, 0.107) | 0.972 (0.959, 0.986) | -0.003 (-0.004, -0.001) |
| 5-year | 0.135 (0.134, 0.137) | 0.138 (0.138, 0.138) | 0.981 (0.969, 0.994) | -0.003 (-0.004, -0.001) |

<sup>1</sup>ACEI, angiotensin-converting enzyme inhibitor; ARB, angiotensin-II receptor blocker; CI, confidence interval; IP, inverse probability.
<sup>2</sup>Dementia-free death represents death without having dementia and treating dementia as the competing event. Results on this outcome were reported to facilitate transparent and comprehensive interpretations of the treatment effects on dementia.

### Secondary Outcomes

During follow-up, 457,605 Veterans experienced dementia or death (39.7/1000 PY), including 43,954 ARB initiators (39.6/1000 PY) and 413,651 ACEI initiators (39.7/1000 PY). ITT effects (RRs and RDs) on this composite outcome paralleled those on dementia outcome alone, with smaller RRs and larger RDs compared to dementia alone. A total of 345,770 Veterans died (28.9/1000 PY), including 33,843 ARB initiators (29.5/1000 PY) and 311,927 ACEI initiators (28.8/1000 PY). Effect sizes and their trends over time for all-cause death were similar to those for dementia-free death (**Table 3**).

Compared with the primary NLP-based dementia outcome, analyses for dementia identified using ICD codes with and without anti-dementia medication use showed similar overall patterns in ITT effects. ICD-only dementia yielded larger relative but smaller absolute effects than the NLP-defined dementia (**eFigure 4** and **eTable 8**). Effects were smaller on both scales when dementia was identified by ICD codes plus medication use. (**eFigure 5** and **eTable 9**).

### Subgroup Analyses

At 5 years, RRs for the primary NLP-based dementia outcome were consistent across most subgroups, with three exceptions. ARB versus ACEI initiation appeared to result in lower dementia risk for individuals aged ≥60 years but not <60 (P-for-interaction = .009), accompanied by a slightly higher RR of dementia-free death in older adults (**Figure 2**).

**Figure 2.**
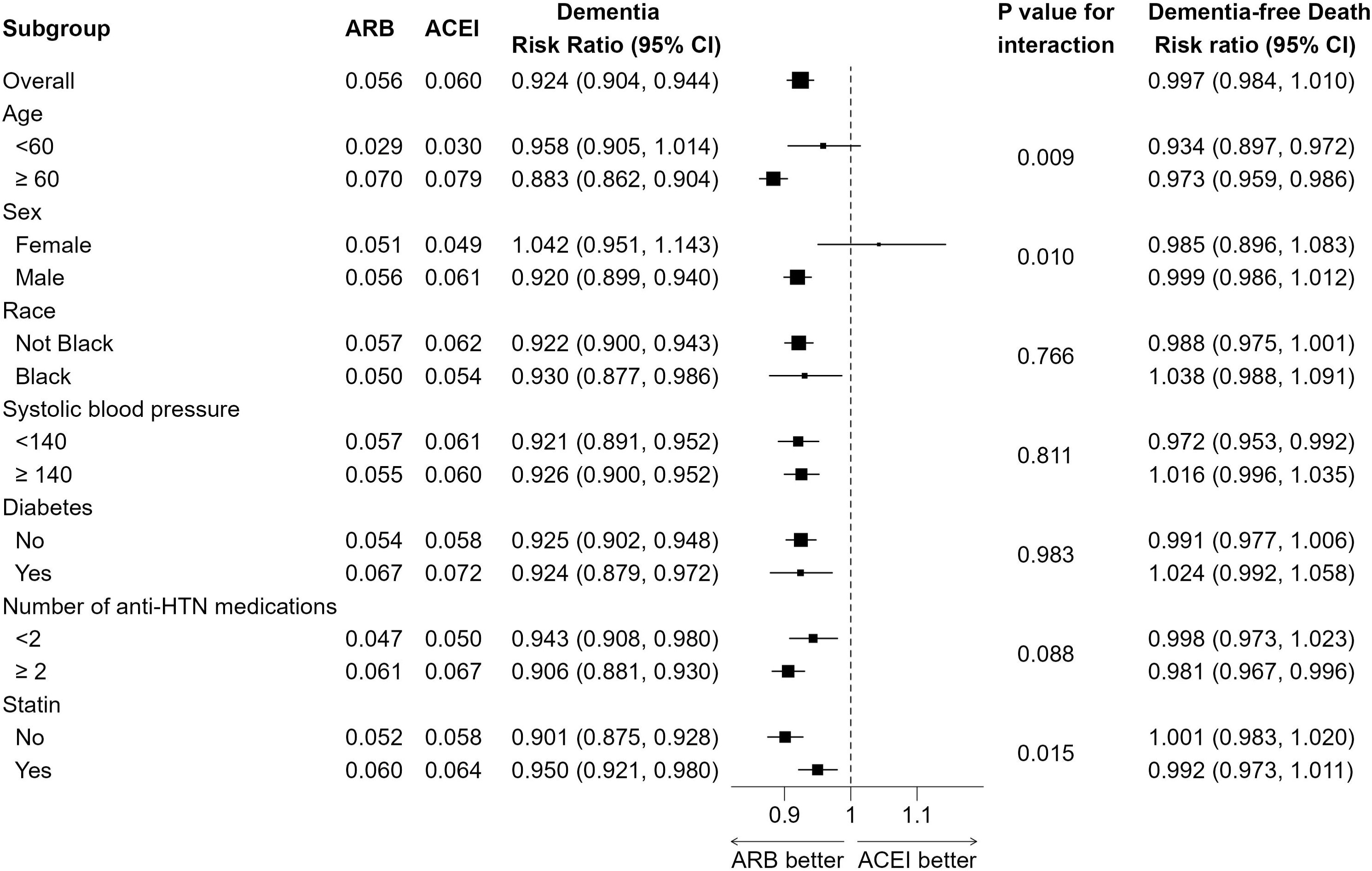
Five-Year Inverse Probability Weighted Effects of ARB Initiation on NLP-based Dementia across Subgroups. The first five columns display absolute risks of dementia under ARB and ACEI initiation, the risk ratio (RR), and the P value for interaction from the Wald test for dementia, indicating whether treatment effects differed across subgroups. Larger squares denote subgroups with larger sample sizes, and error bars represent 95% confidence intervals. The final column presents the RR for dementia-free death.

ARB initiation appeared beneficial in males but not females (P-for-interaction = .010) and more beneficial in non-statin users than in statin users (P-for-interaction = .015), with no evidence of differences in dementia-free death for either comparison. Subgroup cause-specific HRs attenuated over time (**eTable 6**). Results for secondary dementia outcomes are shown in **eFigures 7 and 8**.

### Sensitivity Analyses

Using overlap weighting, ITT effects on both primary and secondary outcomes were larger, with RRs on dementia ranging from 0.83 at 6 months to 0.89 at 5 years and RRs on dementia-free death from 0.84 to 0.93 (**eTable 10; eFigures 9**). Similar patterns were observed for dementia defined by ICD codes with and without anti-dementia medication use (**eTables 11** and **12; eFigures 10 and 11**). In the overlap population, the cause-specific HR for NLP-based dementia decreased over time (**eTable 7**), and subgroup analyses identified similar effect modifiers as in the primary analysis but with larger effect estimates (**eFigure 6** and **eTable 7**).

## DISCUSSION

In this target trial emulation of 2.5 million US Veterans with incident hypertension (95% male), initiation of ARB versus ACEI showed a modestly lower cumulative incidence of dementia, with relative effects attenuating and risk differences increasing over 5 years, while dementia-free death risk was comparable between treatment groups. We observed small but favorable effects with ARB initiation on the composite endpoint of dementia or death and even smaller effects on all-cause mortality over 5 years. Effects on secondary outcomes were similar to those for primary outcomes. Greater protective dementia effects were observed in older and male Veterans and non-statin users, with similar effects on dementia-free death.

Given these results, the lower dementia risk among ARB initiators is unlikely explained by a higher competing risk of death from non-dementia causes. However, given high mortality rate, the modest relative reduction in observed dementia incidence does not necessary imply that ARB initiation slowed the underlying cognitive decline leading to dementia. Our analyses cannot exclude the possibility that dementia-free deaths occurred disproportionately among individuals at higher risk of dementia in the ARB group, which could artifactually lower dementia incidence. Although no biologically plausible mechanism for such selective mortality is known, it cannot be excluded without possibly longitudinal cognitive measurements. While the early effects were surprising, the subsequent attenuation of relative effects may reflect differential adherence rates over time between treatment groups.

Our analyses of the composite endpoint of dementia or death pragmatically assess whether initiating ARBs versus ACEIs improves dementia-free survival and are not affected by complications of competing risks. The small protective effects with ARB initiation make it difficult to rule out uncontrolled confounding, though our study design mitigates this concern. While a 0.5% increase in dementia-free survival is relatively small, it translates to substantial population-level impact (e.g., preventing 50,000 composite events in a population of 10 million).

Our analysis included patients who initiated ARBs or ACEIs between 2000 and 2017. ARB use was low early on but increased in 2010 with the introduction of generic losartan. This pattern of use could introduce bias from secular trends. To address this concern, we conducted sensitivity analyses using overlap weighting, emphasizing individuals with the greatest treatment equipoise. These analyses showed greater effects of ARBs on all outcomes.

Experimental evidence supports a biological rationale for our findings: ARBs selectively block AT1 receptors while maintaining angiotensin II signaling through AT2 and AT4 receptors, pathways linked to reduced neuroinflammation, improved cerebral perfusion, and lower oxidative stress. In contrast, ACEIs lower overall angiotensin II levels, limiting activation of these potentially neuroprotective receptors. Our results are consistent with this proposed mechanism.

Our analyses align with prior findings. A secondary analysis of 2,040 participants from the Systolic Blood Pressure Intervention Trial (SPRINT) found that new users of ARBs versus ACEIs had lower rates of adjudicated amnestic mild cognitive impairment or probable dementia in the standard SBP-lowering arm but not in the intensive arm.^5^ Analyses of large population-based cohorts, including VA, Optum Clinformatics, and multinational databases, report modest reductions in dementia risk of 8-20% among ARB users versus ACEI users.^20,22,44^ A small randomized trial of candesartan versus lisinopril, further suggest that ARBs may preserve executive function or delay cognitive decline.^45^ Our finding of larger protective effects in male Veterans is consistent with a prior study using claims data.^46^ Together, experimental, clinical, and population evidence supports the plausibility that ARBs may provide modest but clinically relevant neuroprotection beyond blood pressure lowering.

Our study has several strengths supporting its validity. The large VHA EHR cohort provided sufficient power to estimate overall ITT effects on incident dementia and to assess treatment effect heterogeneity across key subgroups. Because up to 50% of dementia cases may go undiagnosed in routine care,^47^ we developed and validated an NLP algorithm for identifying non-subtype specific cases from clinical notes. We also evaluated two alternatives: using ICD codes alone and using ICD codes plus anti-dementia medication. Relative to our NLP approach, diagnosis codes alone tended to under-diagnose dementia, though the findings were similar to those based on NLP. Identifying dementia through medication use increases misclassification rates, likely due to off-label prescribing, as noted in the Danish cohort study,^48^ explaining the mixed results observed with this approach.

Methodologically, our study applied several design and analytic advances to strengthen causal inference. The target trial emulation framework and new-user design aligned eligibility, treatment initiation, and start of follow-up (time zero), minimizing selection bias and immortal time bias. An active comparator design further reduced confounding by indication, leveraging the comparable clinical use of ARBs and ACEIs. The unexpected early effects may reflect uncontrolled confounding. Although we adjusted for a comprehensive set of baseline socioeconomic and demographic factors, comorbidities, medication use, and laboratory measures, we could not account for education, socioeconomic status, and baseline cognitive function, both known dementia risk factors; however, income and history of depression were included as reasonable proxies.

Several future directions could strengthen and extend our findings. First, per-protocol analyses could assess the effects of sustained ARB versus ACEI use and clarify whether observed attenuation effects reflect treatment nonadherence. Second, longitudinal dementia-related biomarkers (e.g., plasma tau phosphorylated at threonine 217) could clarify effects on dementia development.^49,50^ Third, comparative effectiveness analyses of other antihypertensive classes and combination versus monotherapy regimens could broaden understanding of cognitive impacts of blood pressure-lowering therapies. Finally, replication in external datasets with greater representation of women and non-Veterans is essential to assess the generalizability of our findings.

## CONCLUSION

Among US Veterans with newly diagnosed hypertension, initiating ARBs versus ACEIs was associated with a modestly lower risk of dementia and a similar risk of dementia-free death. Notably, older and male Veterans, and non-statin users appeared to derive greater benefit from ARBs. Future work is warranted to examine whether sustained ARB versus ACEI use further reduces long-term dementia risk and whether these findings extend to broader demographic and clinical settings.

## Supporting information

Supplementary Online Content

## Data Availability

The Veterans Health Administration (VHA) data used in our study will not be shared due to data sharing restrictions at VHA.

## Acknowledgements

We used the ChatGPT Version 5.2 by OpenAI in December 2025 and January 2026. We used them for editing of author generated content to improve the clarity of the manuscript. The authors take responsibility for the integrity of the content generated.

