## Supplementary Online Content for "Effect of initiating an ARB- versus ACEI-based regimen on dementia risk, a target trial emulation of 2.5 million US Veterans"

### Supplemental Methods

#### Assumptions in emulating the hypothetical trial

By emulating the hypothetical trial specified in the main text, we aligned the eligibility time, treatment initiation time, and the start of follow-up (i.e., time zero) for each patient in the analysis. This alignment ensured that the emulated trial adhered to the principles of the target trial, thereby mitigating selection bias and immortal time bias. Below, we describe the key design choices and assumptions made through the emulation process:

1. **Exclusion of individuals initiating both treatments**.
   We excluded Veterans who initiated both angiotensin-converting enzyme inhibitors (ACEIs) and angiotensin II receptor blockers (ARBs) within 30 days after the index date, as these individuals could not be unambiguously assigned to a treatment group. Given the small number of such individuals (<1%), we considered this exclusion unlikely to introduce meaningful selection bias.
2. **Emulating trial randomization and addressing confounding by indication**.
   To mimic the randomization of a clinical trial and minimize confounding by indication, we implemented the following strategies:
   i. **Active comparator design**: We used an active comparator design, leveraging the fact that there were no clear clinical reasons or prescribing trends favoring ARBs over ACEIs. This design provided us a pseudo-randomization scenario, where the distributions of patient baseline characteristics between the two treatment groups were much more similar than those in a placebo-control design before adjustment.
   ii. **Prespecified covariates**: We identified 66 baseline covariates based on expert domain knowledge to address remaining imbalance between treatment groups. These covariates were selected to capture key demographic, clinical, and healthcare utilization factors. We assumed that adjustment for this covariate set was sufficient to control for confounding by indication bias.
3. **Handling missing data**.
   Based on expert input, we assumed a missing-at-random (MAR) mechanism for a subset of baseline covariates, meaning that the probability of missingness depended only on observed variables and not on the unobserved values themselves. Under this assumption, we used multiple imputation by chained equations (MICE) to handle missing data. For the remaining baseline covariates, primarily laboratory values, where the MAR assumption was unlikely to hold, a separate “unknown” category was created.

#### Baseline covariates

The 66 baseline covariates encompassed sociodemographic factors such as age, sex, race and ethnicity, income, insurance type, veterans integrated service network, priority group, homelessness, and smoking status. Clinical measures included body mass index, systolic and diastolic blood pressure, heart rate, total cholesterol, high-density lipoprotein cholesterol, low-density lipoprotein cholesterol, triglycerides, hemoglobin A1c, estimated glomerular filtration rate, albumin-to-creatinine ratio, serum sodium, serum potassium, and ejection fraction. We also incorporated comorbidities including cardiovascular disease, myocardial infarction, peripheral artery disease, stroke, heart failure, cancer, cirrhosis, diabetes, depression, and substance use disorder, among others. For medication variables, we identified use of statins, aspirin, sodium-glucose cotransporter 2 inhibitors, antidepressants, total number of antihypertensives, and non–ACEI/ARB antihypertensive agents (e.g., beta-blockers, calcium channel blockers, diuretics). Measures of health care utilization, such as frequency of primary care visits, rural vs urban setting, academic vs nonacademic setting, and clinical center patient volume, were also included.

Our analysis included patients who initiated ARBs or ACEIs between 2000 and 2017. ARB use was low early on but increased in 2010 with the introduction of generic losartan. This pattern of use could introduce bias from secular trends. To account for this, we also include the year of ARB or ACEI initiation in our propensity score model. Please see **eTable 1** for detailed definitions.

#### Baseline treatment propensity score model

To achieve covariate balance between ACEI and ARB initiators in both the overall cohort and predefined subgroups, we estimated the probability of initiating an ARB using a logistic model that included 66 baseline covariates and their two-way interactions with seven prespecified subgroup variables: age (<60 vs ≥60 years), sex (female vs male), race (Black vs non-Black), systolic blood pressure (<140 vs ≥140 mm Hg), diabetes status, number of antihypertensive agents (<2 vs ≥2), and statin use. Although the large sample size permitted estimation of this complex model, including all interaction terms would substantially increase variance. Therefore, we used the least absolute shrinkage and selection operator (LASSO) to identify important covariate-subgroup interactions while retaining all main effects of baseline covariates. The final logistic model, incorporating the selected interactions and baseline covariates, was refitted to estimate propensity scores (PSs) in the full sample and within each bootstrap sample to account for PS uncertainty. For a given sample, a single PS model was used to estimate average treatment effects in the overall and subgroup analyses, consistent with the principle that one true PS model underlies the relationship between treatment, covariates, and subgroup variables. This approach aligns with the recommendations of Yang et al. (2021).

#### Identifying dementia outcomes using natural language processing

We developed and validated a natural language processing (NLP) approach to identify dementia events from free-text clinical notes within our study population. The process began with iterative sampling to create an annotated dataset, employing multiple strategies to balance annotation efficiency with comprehensive coverage. These strategies included keyword-based sampling (using 96 expert-curated terms), note-type-based sampling, clinical-specialty-based sampling, and vector-similarity-based sampling. Two trained annotators, achieving an inter-annotator agreement greater than 0.93, annotated 2,152 notes from 1,339 patients over 16 rounds, with a third nursing informatist adjudicating discrepancies in overlapping annotations.

The annotated data were divided into training (1,071 patients) and testing (268 patients) sets and converted into sentence-level labels. A multi-label sequence classification model was trained on the training set using a locally pretrained Bidirectional Encoder Representations from Transformers (BERT) model developed by the Veterans Affairs (VABERT), which was originally trained on 3 million notes. Due to hardware constraints and the large corpus size (2.1 billion notes in total), the solution was further optimized using a hybrid framework. This framework combined rule-based filtering, implemented through SQL exclusion logic and 45 high-recall keywords, with a binary support vector machine (SVM) sentence classifier. The SVM model was trained using training data to classify whether a sentence contained any annotation. We used the Term Frequency-Inverse Document Frequency (TfIdf) vectorizer with linear kernel and class-balanced weights due to the highly skewed data distribution. It achieved a precision of 0.76, a recall of 0.93, and an F1 score of 0.84 on the test dataset. The SVM classifier performed a two-step filtering process before applying the trained VABERT model to generate fine-grained, sentence-level labels. A subsequent rule-based component, guided by annotation guideline logic, aggregated the sentence-level labels into document-level and, ultimately, patient-level labels. This hybrid approach achieved a patient-level F1 score of 0.87, precision of 0.90, and recall of 0.84.

#### Handling the competing risk of death

Given that the overall event rate of death was three times higher than the event rate of dementia (depending on the definitions) in our data, we applied a formal competing risk framework to appropriately account for the competing risk of death. We considered a discrete time setting and estimated the cumulative incidence function (CIF) for both dementia and dementia-free death outcomes using inverse probability (IP)-weighted Aalen-Johansen estimator to provide a comprehensive analysis:

1. The cumulative incidence of dementia (denoted as ‘DA’), treating death as a competing event, i.e., $\mathrm{CIF}_{\mathrm{DA}}\left( t \right)=\sum_{k:t_{k}\leq t} I_{\mathrm{DA}}(t_{k})=\sum_{k:t_{k}\leq t} S\left( t_{k}-1 \right)\lambda_{\mathrm{DA}}^{\mathrm{cs}}\left( t_{k} \right)$
2. The cumulative incidence of death (denoted as ‘DE’), treating dementia as a competing event, i.e., $\mathrm{CIF}_{\mathrm{DE}}\left( t \right)=\sum_{k:t_{k}\leq t} I_{\mathrm{DE}}\left( t_{k} \right)=\sum_{k:t_{k}\leq t} S\left( t_{k}-1 \right)\lambda_{\mathrm{DE}}^{\mathrm{cs}}\left( t_{k} \right)$,

where $I(t_{k})$ indicates the incidence of DA or DE at time $t_{k}$, $S\left( t_{k}-1 \right)$ represents the IP-weighted overall probability of surviving both events at time $t_{k}-1$, and $\lambda^{\mathrm{cs}}\left( t_{k} \right)$ denotes the IP-weighted cause-specific hazard, defined as the instantaneous rate of the event (e.g., ‘DA’) at time $t_{k}$ given that a subject has survived up to $t_{k}$ without any events. It is obvious that the sum of the two cumulative incidences at any time $t_{k}$ equals to the cumulative incidence (i.e., risk) of the composite outcome of dementia or death, which was analyzed as a secondary outcome in our work.

These two sets of CIFs should be interpreted jointly to assess whether a lower dementia risk in one treatment group may be explained by a higher competing risk of death. Subdistribution CIFs for the two outcomes thus provide a more transparent and comprehensive depiction of causal effects. Unlike cause-specific hazard models, which censor individuals at death and assume independent censoring, the CIF approach incorporates the overall survival function for both dementia and death events, thereby avoiding this assumption.

#### Overlap weighting

While decent common support between ARB and ACEI initiators in our study, we observed limited overlap in the region where estimated propensity scores (PS) exceeded 0.25 (**eFigure 2**). This indicated that few ACEI initiators had a high probability of receiving ARBs. In addition, the limited use of ARB before Iosartan became generic in 2010 may further exacerbate the nonoverlap issue.

To account for nonoverlap on our intention-to-treat effect estimates, we performed sensitivity analyses using overlap weighting, with weights $W_{i}$ defined as:

$$W_{i}=\left\{ \begin{aligned} \hat{e}_{i}, A_{i}=ACEI; \\ 1-\hat{e}_{i}, A_{i}=ARB, \end{aligned} \right.$$

where $A_{i}$ indicates the treatment that Veterans initiated (ACEI is the reference group) and $\hat{e}_{i}$ is the estimated PS. Note that in our study where ARBs were used roughly one-tenth as often as ACEIs, the average treatment effect in the overlap population approximates the “average treatment effect in the treated” (ATT).

Unlike the IP of treatment weighting approach that estimates the average treatment effects (ATEs) in the entire population, overlap weighting targets a distinct causal estimand: the ATE in the overlap population (ATO). This population comprises Veterans with the most treatment equipoise -- those with the most overlap in observed characteristics between treatment groups. Some argue that ATO is a closer estimand to the treatment effect estimand in randomized control trials than ATEs. The overlap weighting approach provides two key advantages: (1) it inherently achieves exact covariate balance between treatment groups when the PS is estimated using a logistic model, and (2) it eliminates the need for arbitrary decisions regarding weight truncation or trimming that are often required with IP weighting. This sensitivity analysis allowed us to assess whether our primary findings were robust when focusing specifically on the overlap subpopulation.

**Interpretation of treatment effect estimates under competing risk**

Under the competing risk framework, the incidence of dementia at time t represents the proportion of individuals who develop dementia among those still at risk at time t-1, where “at risk” refers to being free of both dementia and death. The probability of remaining at risk is the dementia-free survival, which is the survival probability of the composite outcome of dementia or death.

A treatment may appear to reduce the risk of dementia even when the true effect is null or harmful if the “at risk” population is substantially smaller in that group, for example, if more patients died before developing dementia. Because the size of the “at risk” set directly influences the number of observed dementia events (the numerator), interpreting treatment effects under a competing risk framework requires careful evaluation of differences in “at risk” rates between treatment groups. Particular caution is warranted when one group shows a lower risk of dementia but a higher risk of the composite outcome of dementia or death, or vice versa.

### Supplemental Figures

#### eFigure 1. Flowchart of Cohort Identification. Flowchart illustrating the inclusion and exclusion criteria used to identify new ARB or ACEI initiators within the VHA database.

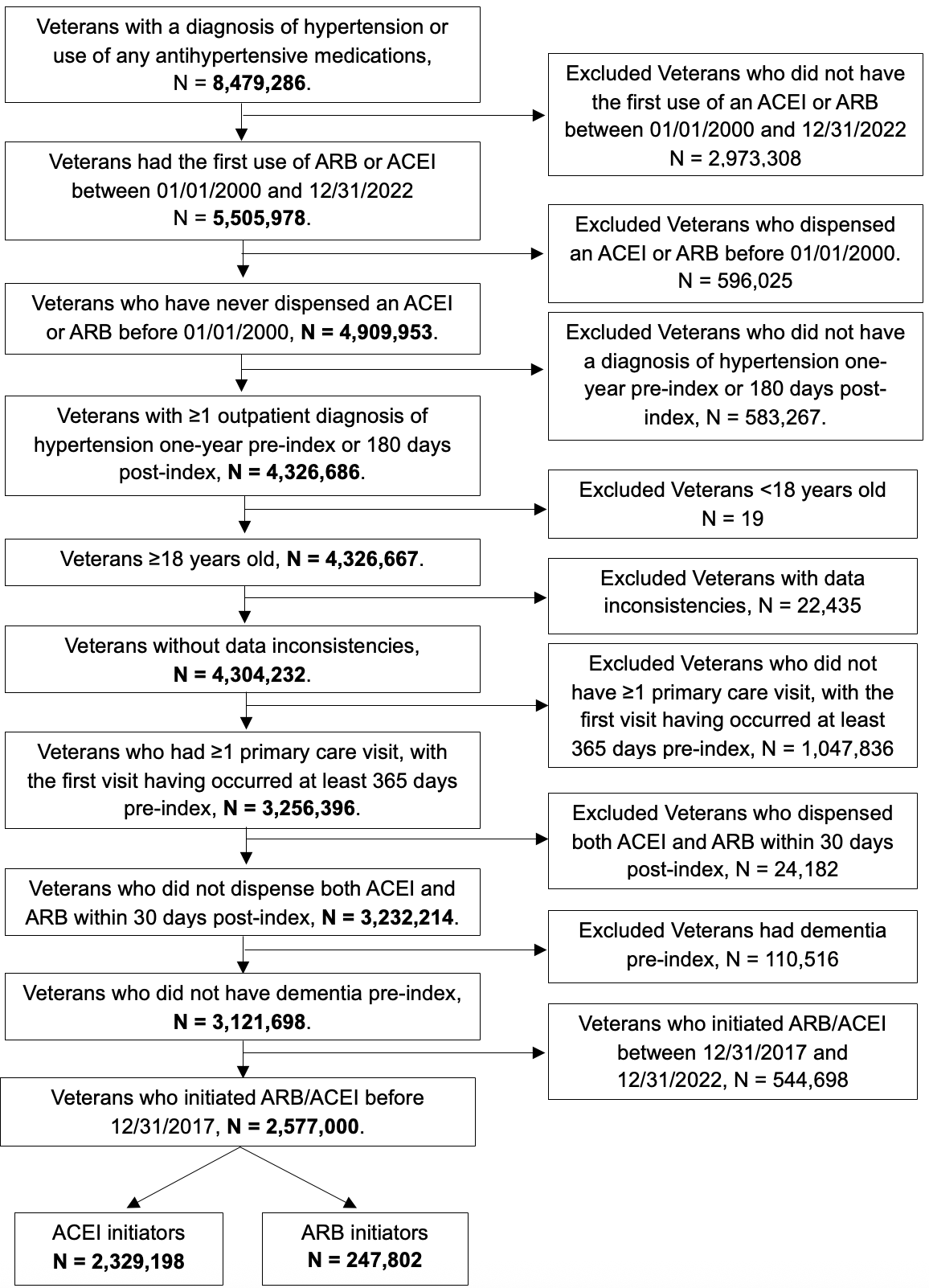

#### eFigure 2. Propensity Score Overlap Before Inverse Probability Weighting. Distribution of propensity scores, showing overlap between treatment groups.

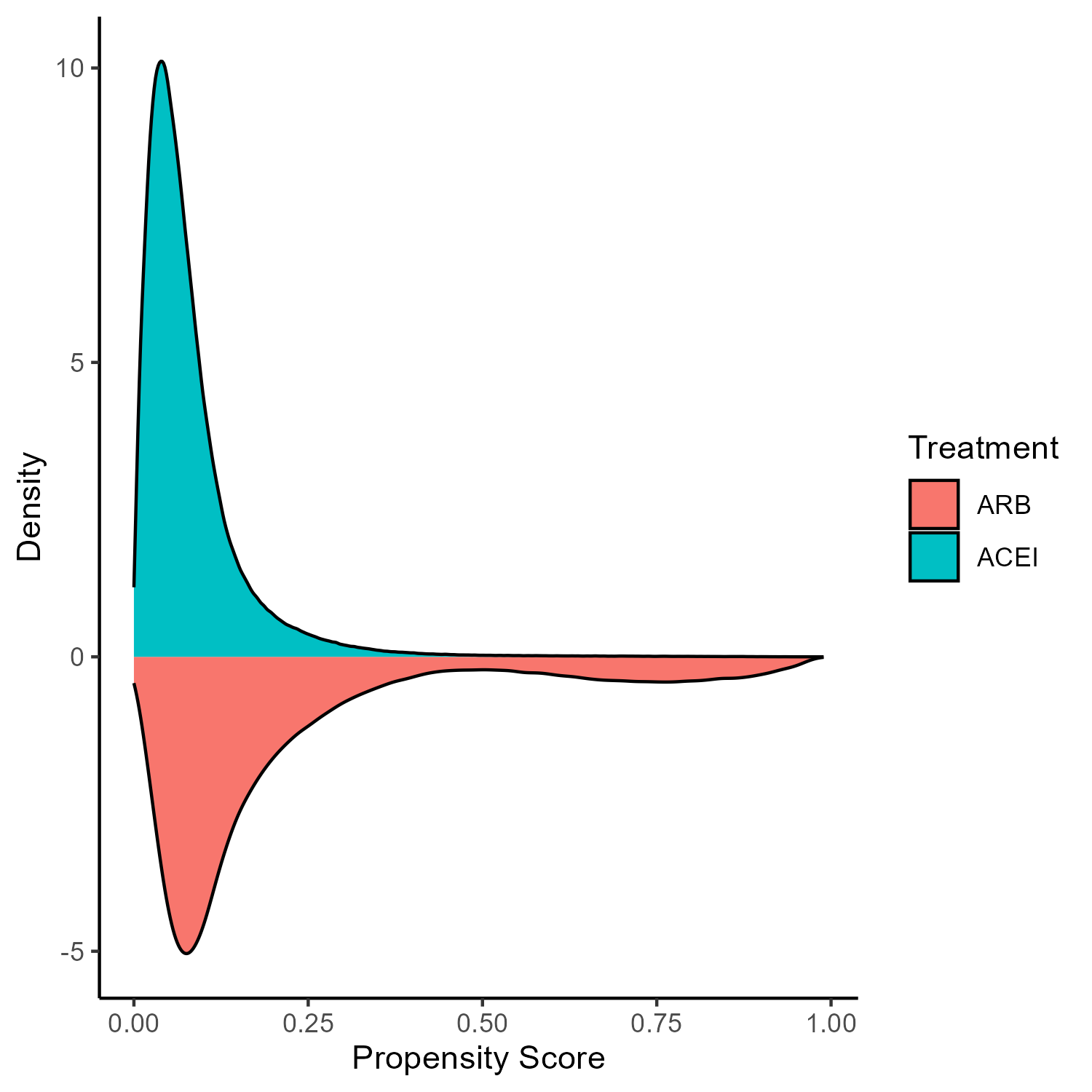

#### eFigure 3. Covariate Balance Before and After Inverse Probability Weighting. Absolute standardized mean differences for baseline covariates before and after weighting, indicating improved balance across treatment groups.

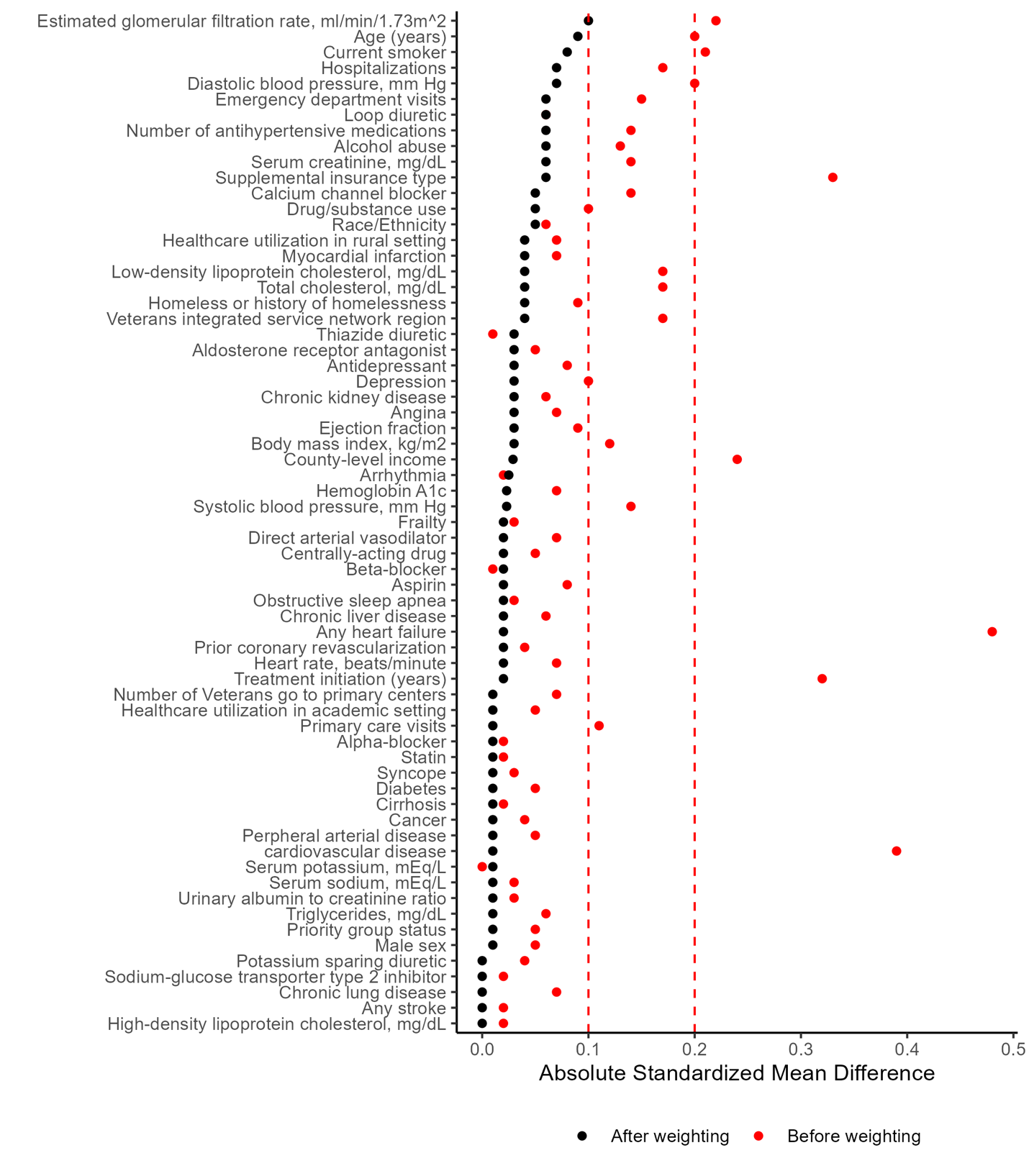

#### eFigure 4. Inverse Probability Weighted Cumulative Incidence of Dementia and Dementia-free Death Identified Using Diagnosis Codes Only.

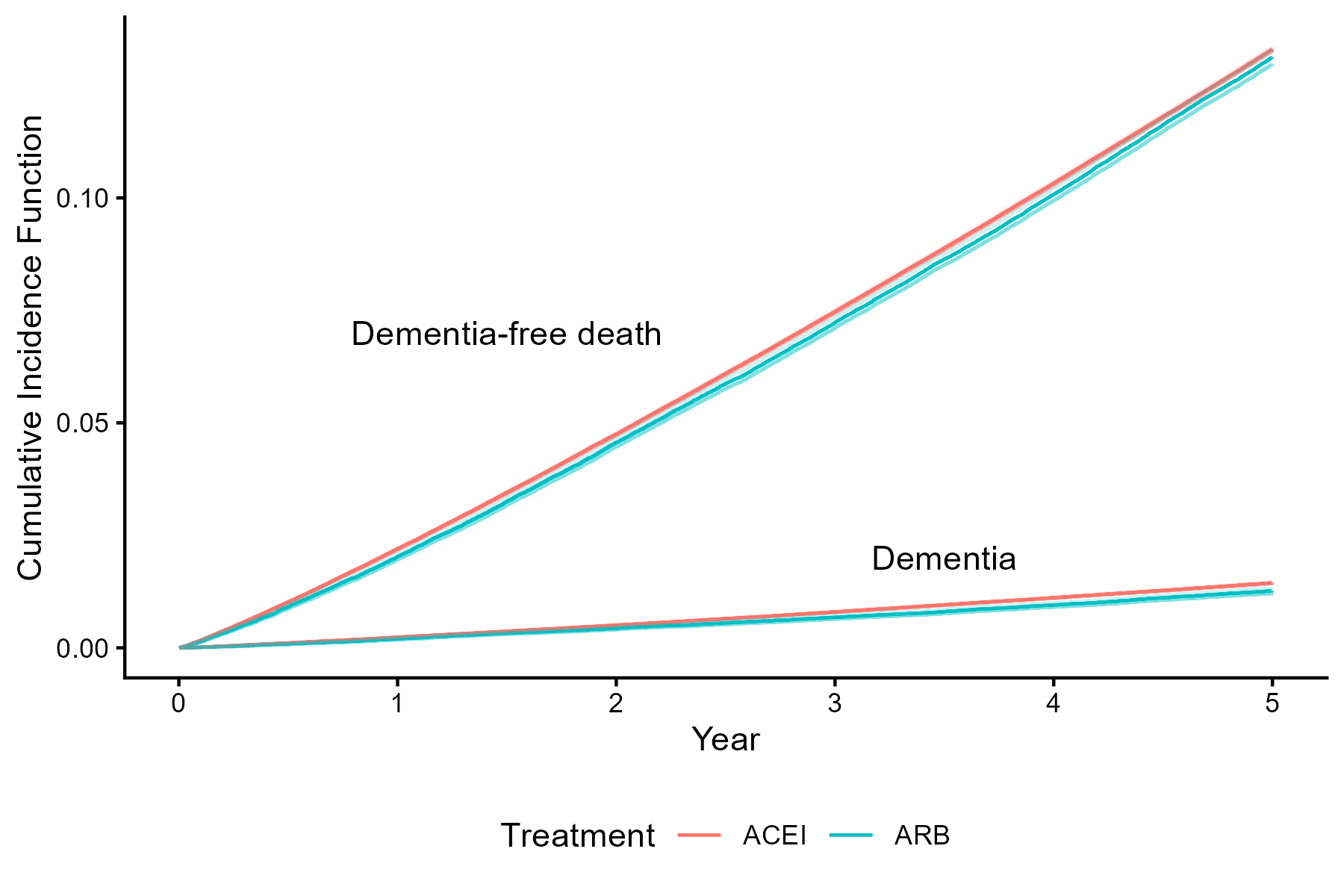

#### eFigure 5. Inverse Probability Weighted Cumulative Incidence of Dementia and Dementia-free Death Identified Using Diagnosis Codes and Anti-dementia medication use.

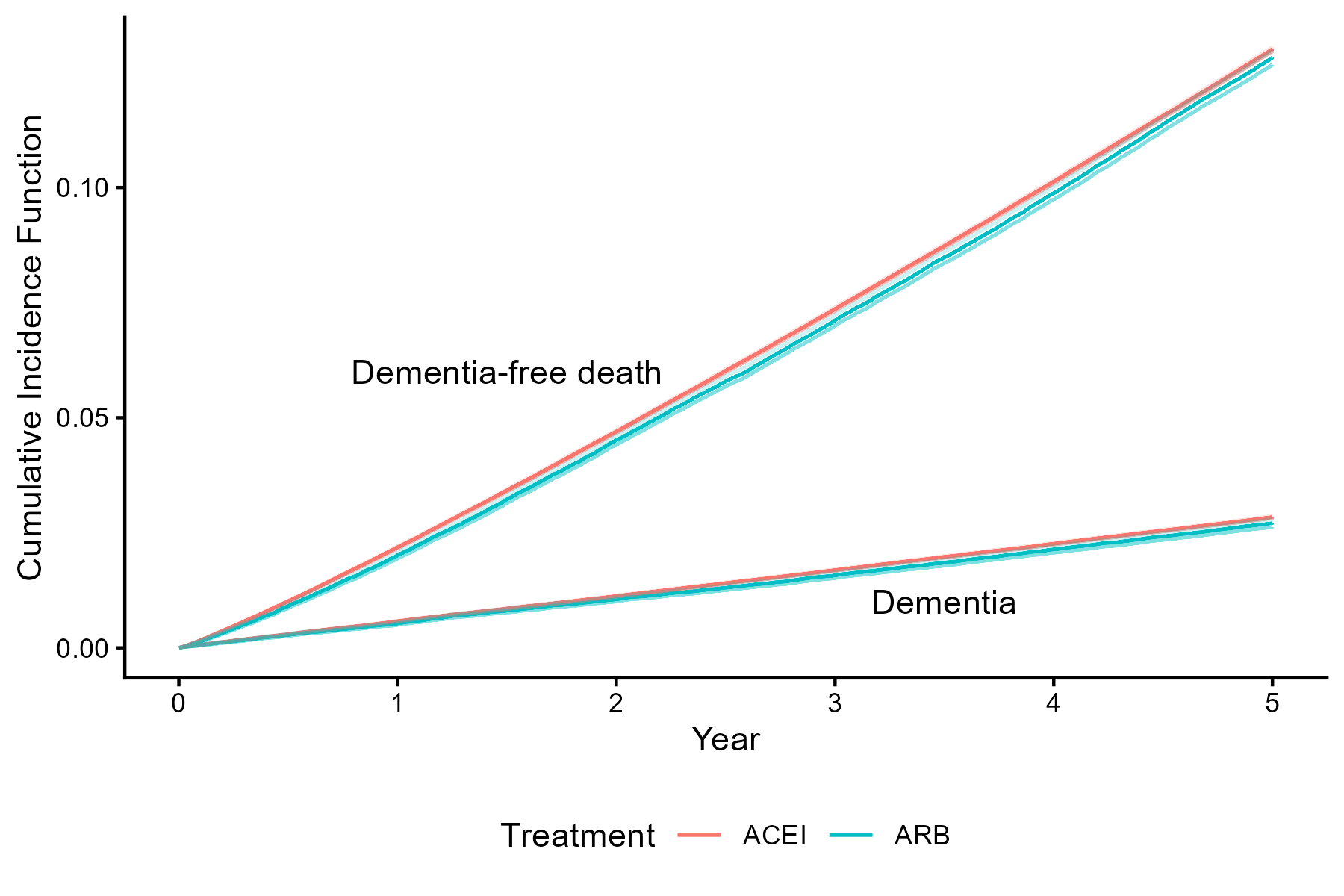

#### eFigure 6. Five-Year Overlap Weighted Effects of ARB Initiation on NLP-based Dementia across Subgroups. The first five columns display absolute risks of dementia under ARB and ACEI initiation, the risk ratio (RR), and the P value for interaction for RR from the Wald test for dementia, indicating whether treatment effects differed across subgroups. Larger squares denote subgroups with larger sample sizes, and error bars represent 95% confidence intervals. The final column presents the RR for dementia-free death.

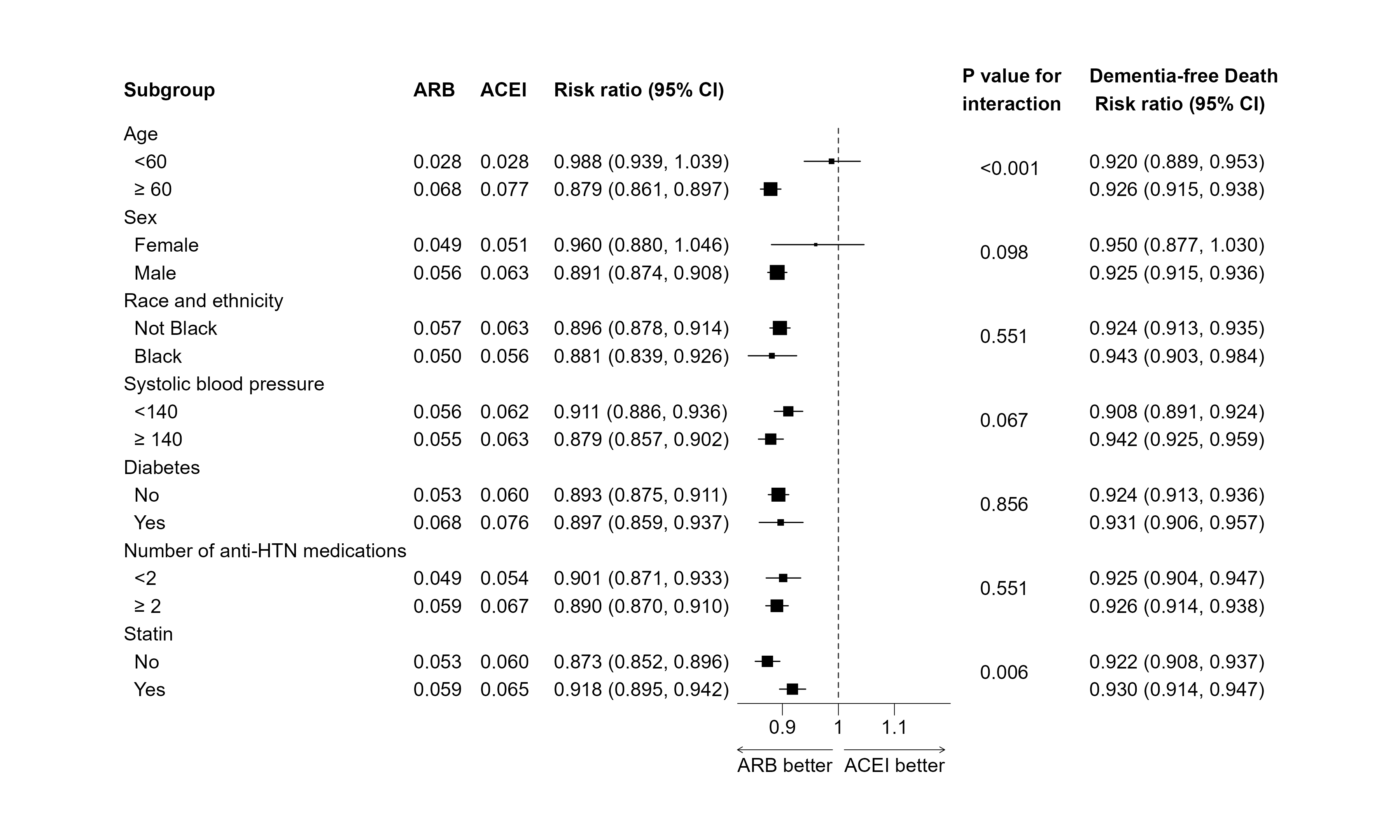

#### eFigure 7. Five-Year Inverse Probability Weighted Effects of ARB Initiation on ICD-based Dementia across Subgroups. The first five columns display absolute risks of dementia under ARB and ACEI initiation, the risk ratio (RR), and the P value for interaction for RR from the Wald test for dementia, indicating whether treatment effects differed across subgroups. Larger squares denote subgroups with larger sample sizes, and error bars represent 95% confidence intervals. The final column presents the RR for dementia-free death.

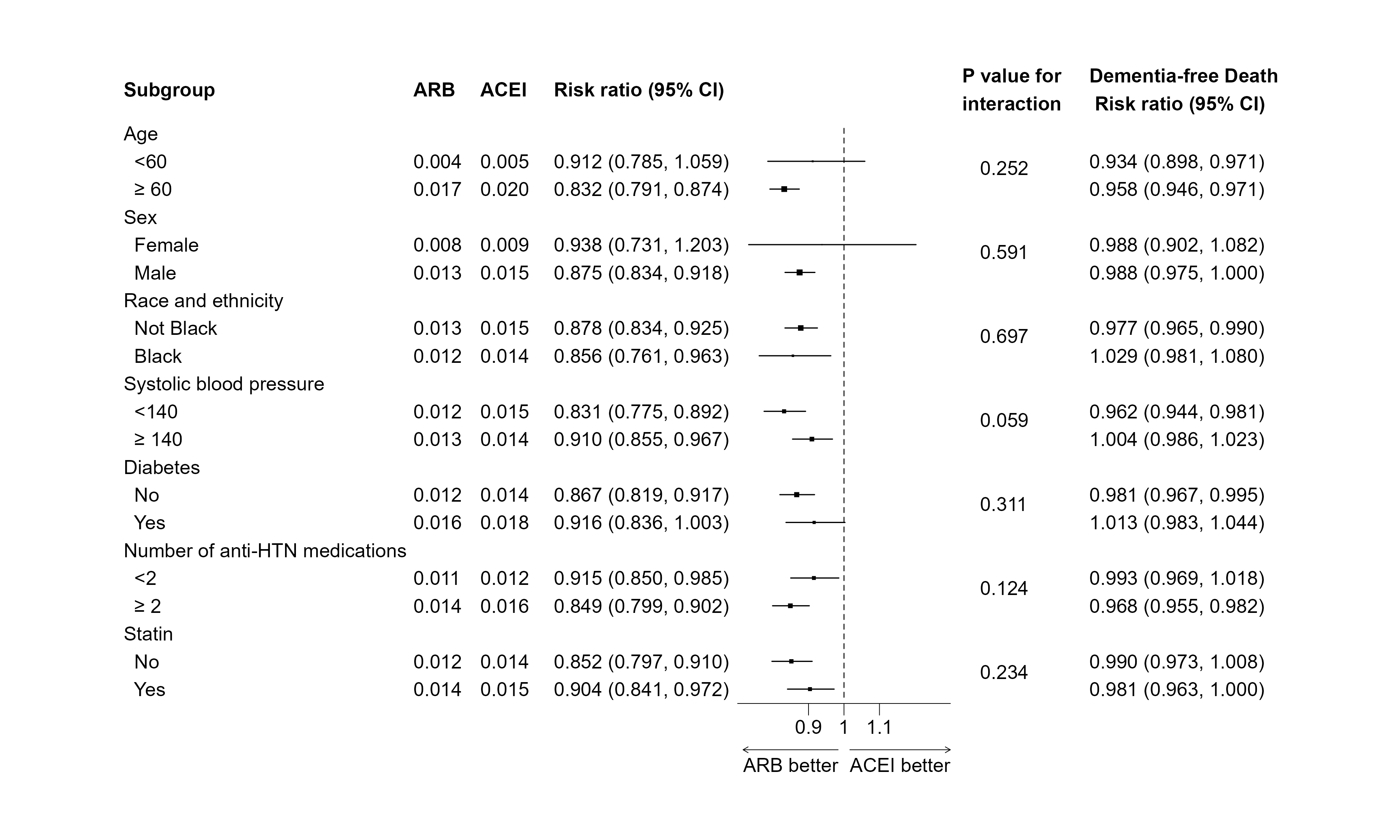

#### eFigure 8. Five-Year Inverse Probability Weighted Effects of ARB Initiation on ICD- and Medication-based Dementia across Subgroups. The first five columns display absolute risks of dementia under ARB and ACEI initiation, the risk ratio (RR), and the P value for interaction for RR from the Wald test for dementia, indicating whether treatment effects differed across subgroups. Larger squares denote subgroups with larger sample sizes, and error bars represent 95% confidence intervals. The final column presents the RR for dementia-free death.

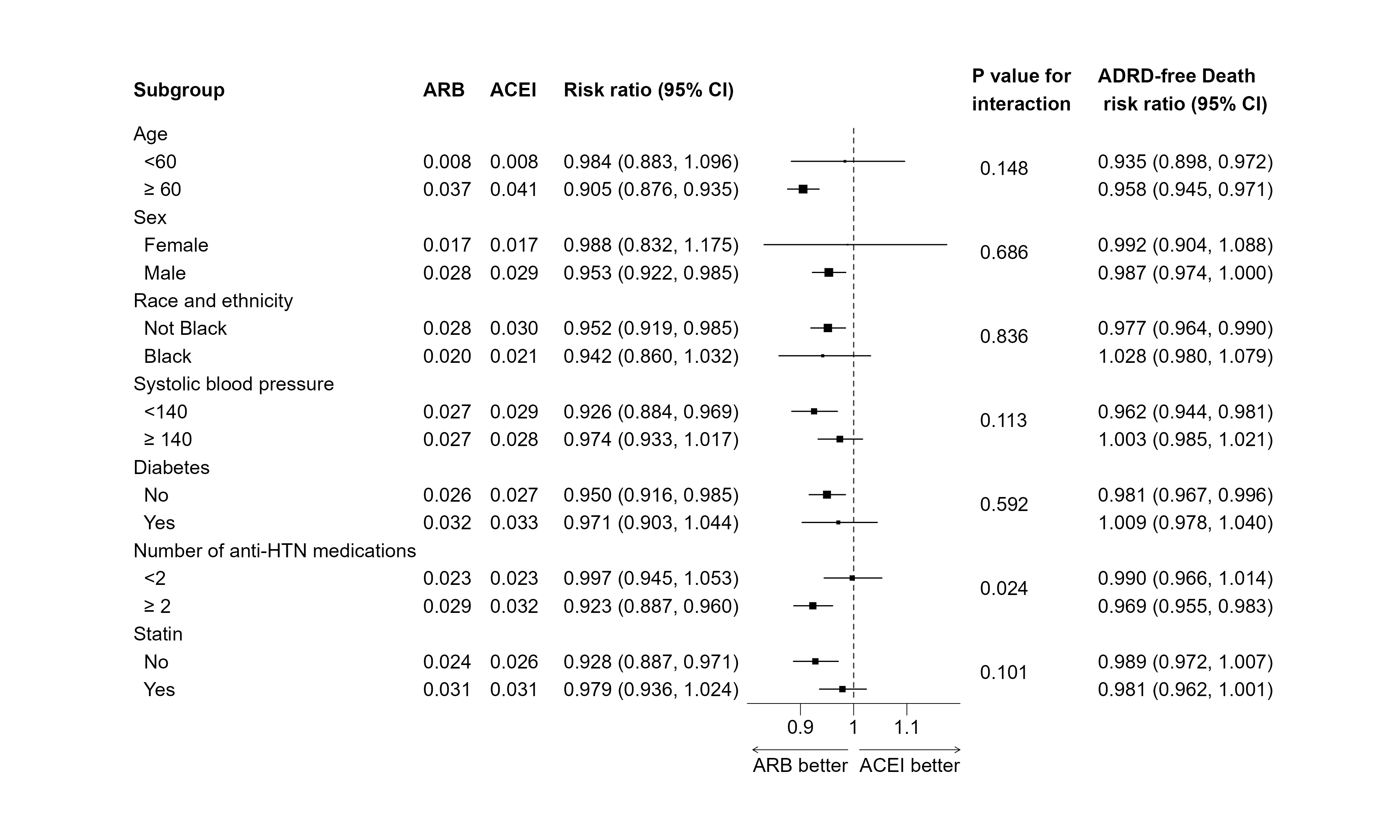

#### eFigure 9. Overlap Weighted Cumulative Incidence of Dementia Identified Using NLP.

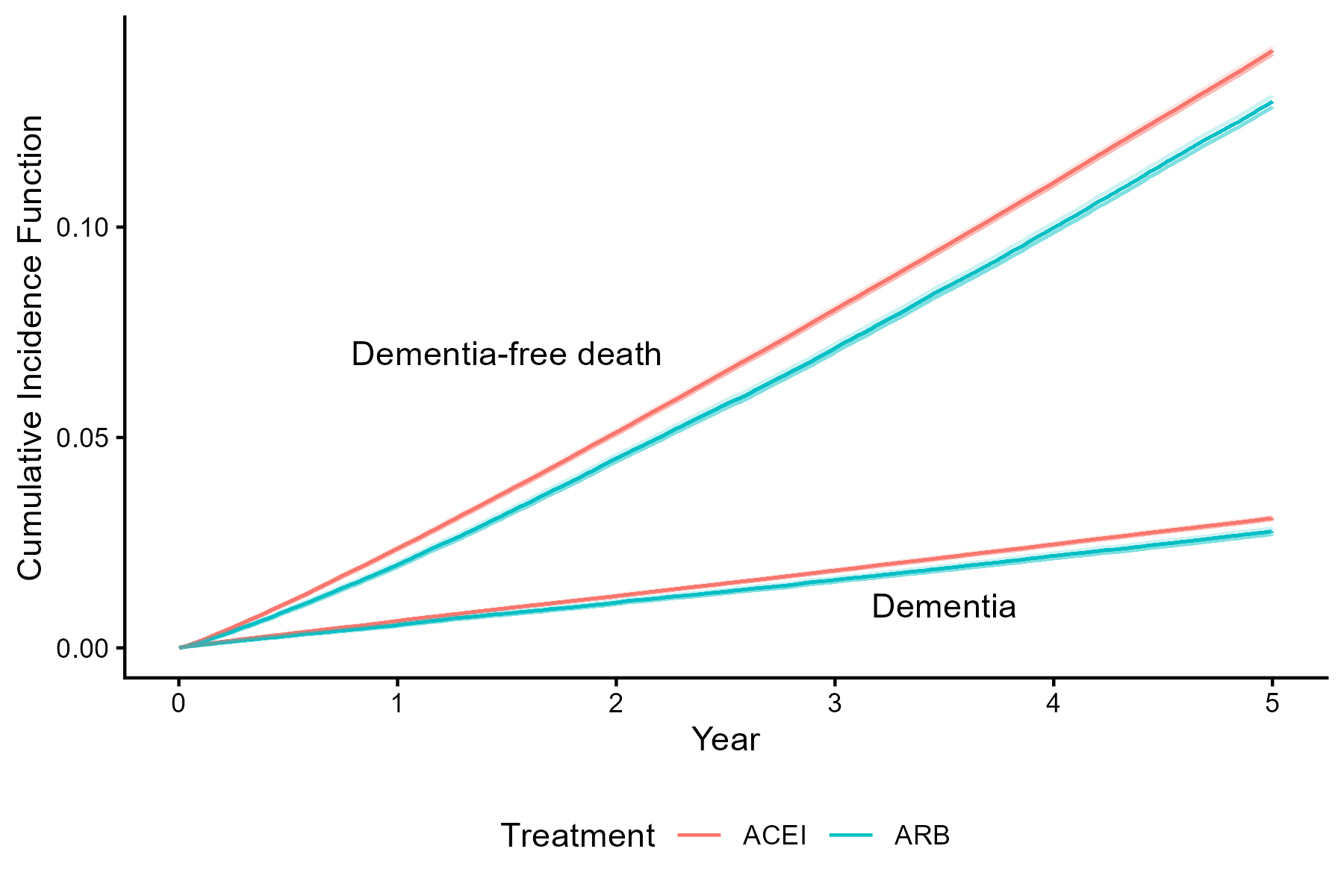

#### eFigure 10. Overlap Weighted Cumulative Incidence of Dementia Identified Using Diagnosis Codes Only.

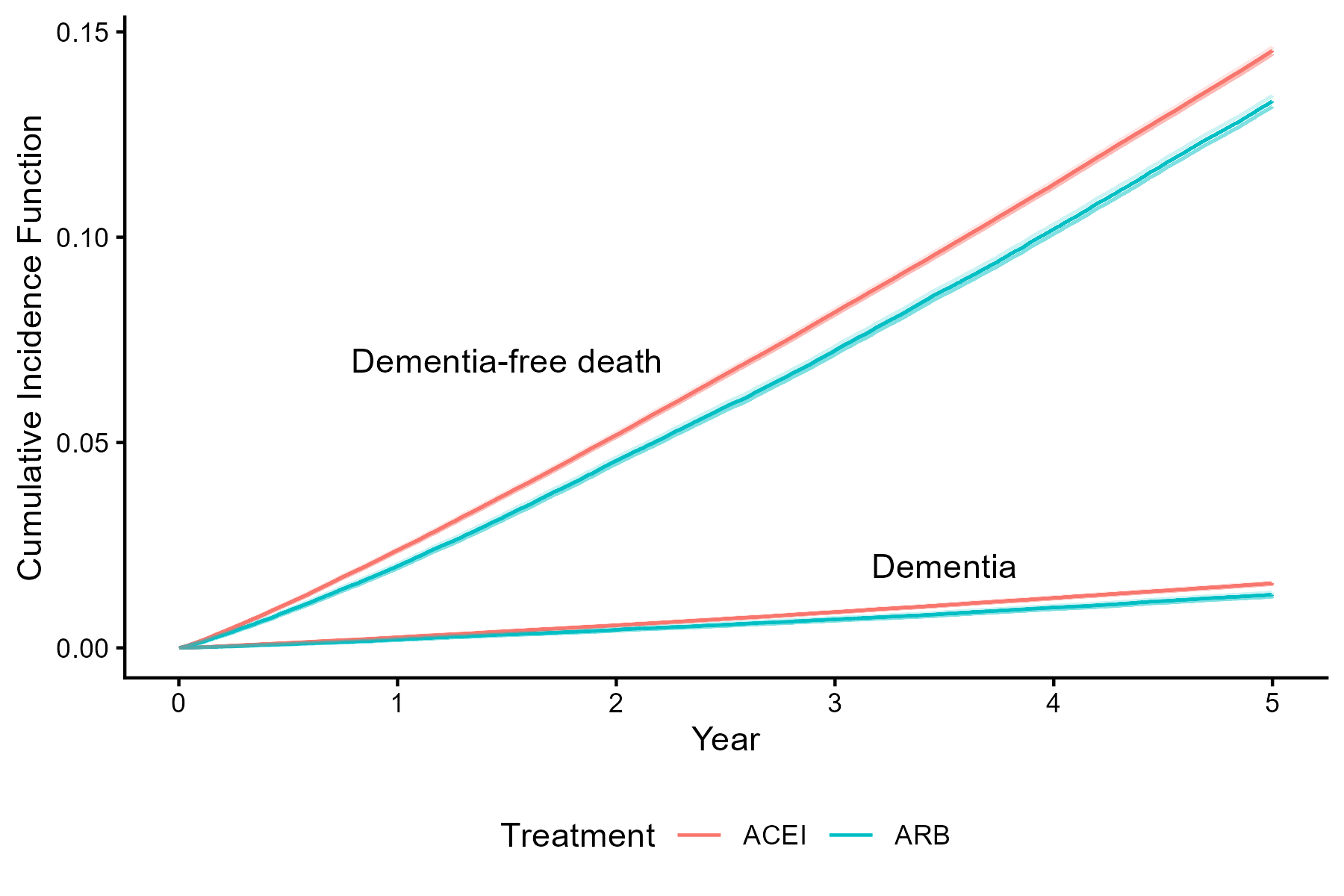

#### eFigure 11. Overlap Weighted Cumulative Incidence of Dementia Identified Using Diagnosis Codes and Anti-dementia Medication Use.

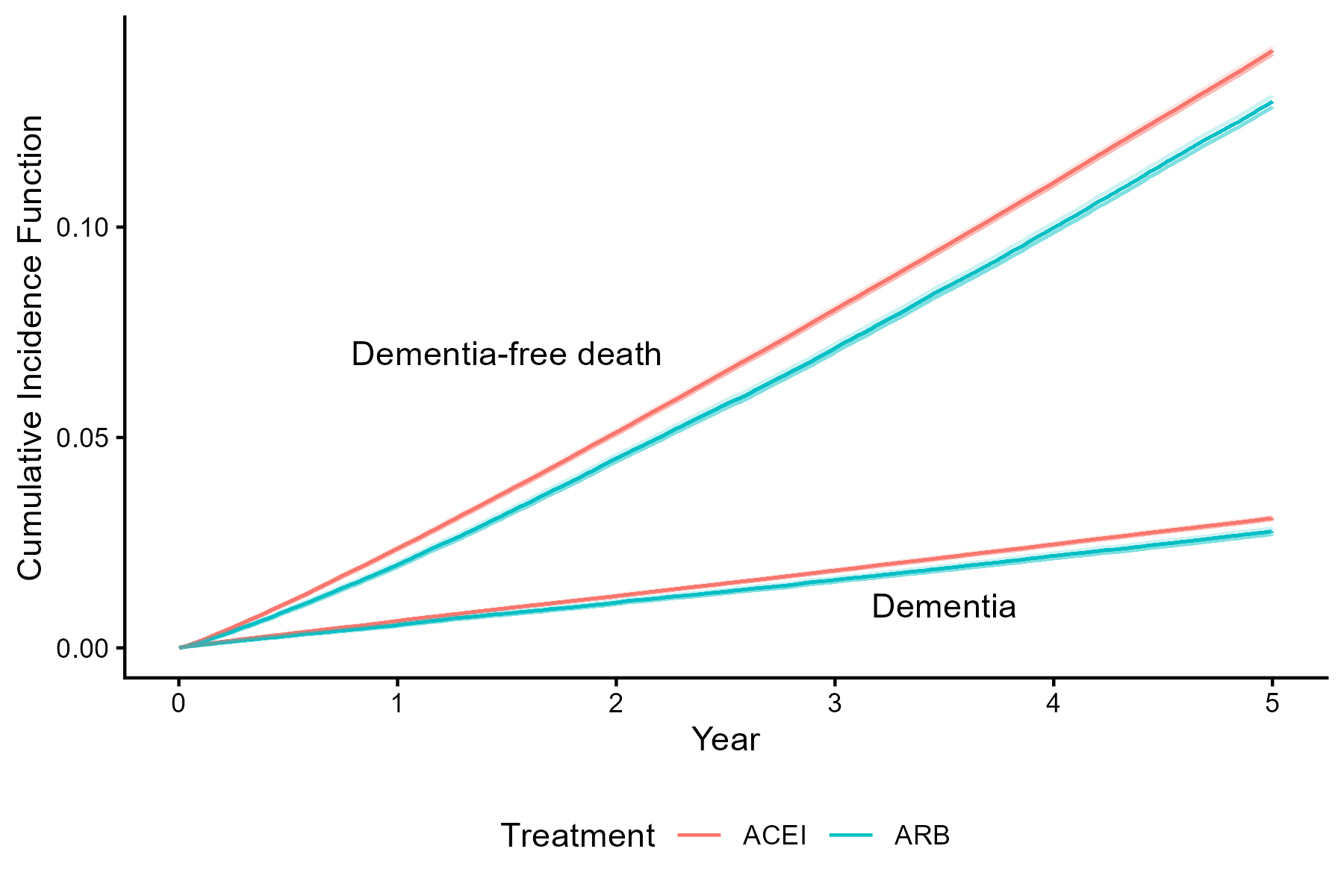

#### eFigure 12. Survival curves for time to deviation from protocol (i.e., nonadherence). A patient is adhered in the current month if s/he used the assigned meds in any of the past 9 months.

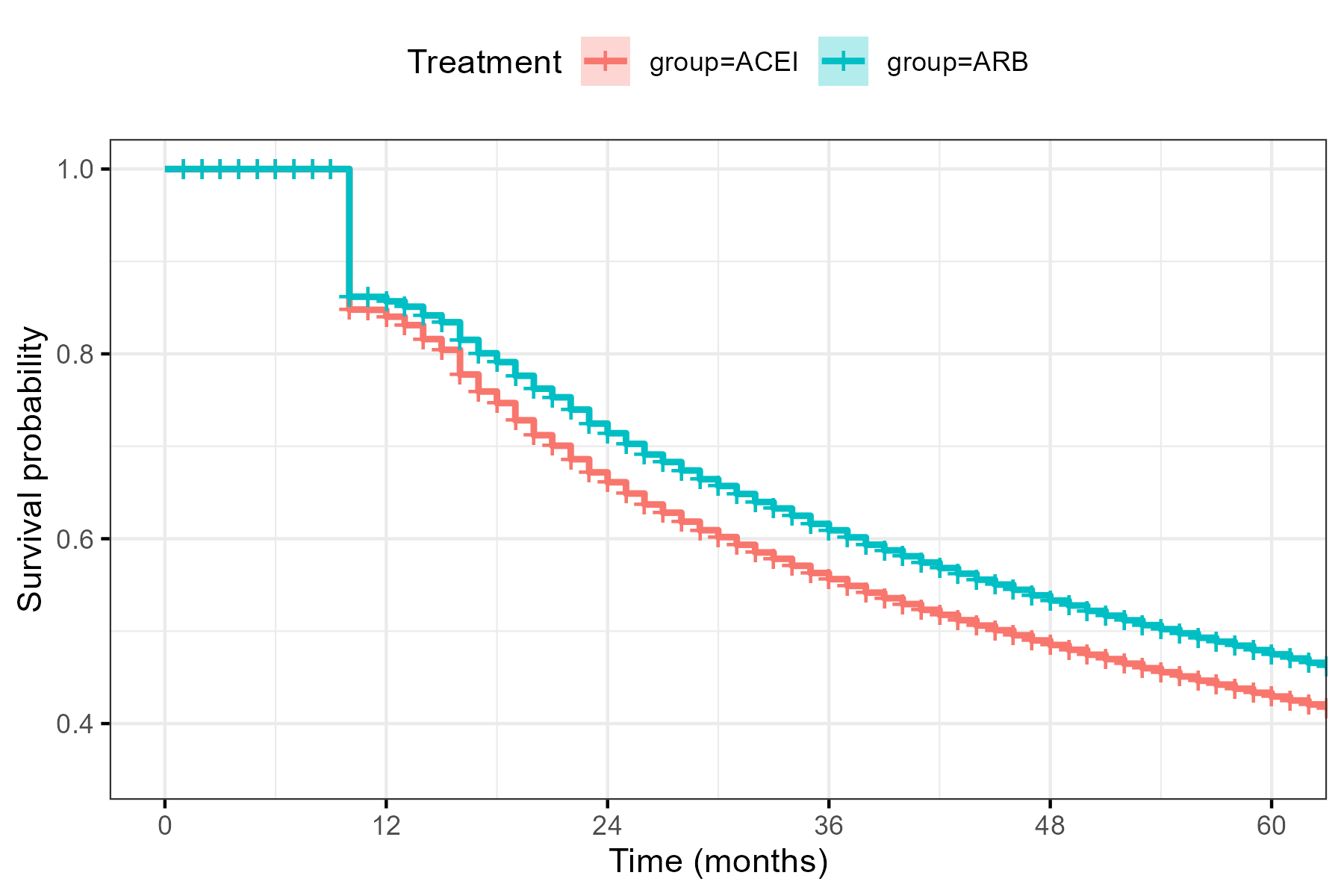

### Supplemental Tables

#### eTable 1. Definitions of Baseline Covariates

| **Variable** | **Definition** |
| --- | --- |
| *Socio-demographics* |  |
| Age | Age of Veterans calculated on the index date based on their date of birth available in the Veteran summary file. |
| Sex | Male or female |
| Race | White, Black, Asian, American Indian or Alaskan Native, Native Hawaiian or Other Pacific Islander, Other, and Missing |
| Ethnicity | Hispanic or non-Hispanic |
| County-level per-capita income | The Veteran’s FIPS code will be matched to the county-level per-capita income available from the US Bureau of Economic Analysis (<https://apps.bea.gov/regional/histdata/index.cfm>). The county-level per-capita income value was also matched to the Veteran’s index year. |
| Veterans Integrated Service Network (VISN) | Defined by receipt of care in one of the 23 VISNs. Will be categorized into 4 regions after data query: Northeast, Southeast, Continental, and Pacific according to the VA regional offices map.^4^ The Northeast region is comprised of VISNs 1, 2, 4, 5, 10, and 12. The Southeast region is comprised of VISNs 6, 7, 8, 9, and 16. The Continental region is comprised of VISNs 15, 17, 18, 19, and 23. Finally, the Pacific region is comprised of VISNs 20, 21, and 22. |
| Supplemental insurance type | Coded as Medicare, Medicaid, private (all insurance external to Medicare/Medicaid), none, and unknown. |
| Priority group status | Coded as 1 through 9 or multiple. |
| Current Smoking | Any of the following within one year prior to the index date (including the index date):   (a) ICD-9 codes:     a. ≥1 hospitalization with a discharge diagnosis code (any position) of tobacco use of 305.1, 649.0x, 989.84, or V15.82) in any discharge position     b. ≥1 physician evaluation and management visit with a diagnosis code (any position) of tobacco use of 305.1, 649.0x, 989.84, or V15.82) in any discharge position (b) ICD-10 codes:     a. ≥1 hospitalization with a discharge diagnosis code (any position) of tobacco use of F17.200, F17.201, F17.210, F17.211, F17.220, F17.221, F17.290, F17.291, or Z87.891) in any discharge position     b. ≥1 outpatient visit with a diagnosis code (any position) of tobacco of F17.200, F17.201, F17.210, F17.211, F17.220, F17.221, F17.290, F17.291, or Z87.891) in any discharge position (c) ≥1 hospitalization with a discharge diagnosis code or physician evaluation and management visit of tobacco use with a CPT code (any position) of 99406, 99407, G0436, G0437, G9016, S9453, S4995, G9276, G9458, 1034F, 4004F, 4001F (d) ≥1 pharmacy claim for nicotine or varenicline in the 379 days before the index date (including the index date). (e) Self-reported smoking status as “current” |
| Homelessness or history of homelessness | Meets at least one of the following criteria using all available claims prior to the index date (including the index date):   a) Claim with at least one of the following outpatient stop codes: 201, 504, 508, 511, 522, 528, 529, 530, 590, 591, 592 b) Outpatient claim with at least one of the following diagnosis codes in any position: V60.0, V60.1, V60.89, V60.9, Z59.0, Z59.1, Z59.8, Z59.9 c) Inpatient claim with at least one of the following treatment specialty codes (SpecialtyIEN): 28, 29, 37, 39 d) Screens as positive for homelessness or at risk for homelessness using the HSCR (homeless screening clinical reminder)^5^   Please note: For future studies in which homelessness is a key variable (ie, social determinants of health), this variable will need to be narrowed/modified. |
| *Vital signs and laboratory measures* |  |
| Body mass index | Body mass index on or closest to the index date during the one-year pre-index period. Height and weight measurements do not need to be on the same day. This variable will be calculated from height and weight observations in the CDW vital status file as weight (in kilograms) divided by height (in meters) squared. Values ≥10 kg/m^2^ and ≤80 kg/m^2^ will be retained; outliers will be coded as missing. |
| Systolic blood pressure_pre6 months | Average of all SBP values in the 6 months prior to the index date (including the index date) corresponding to an outpatient encounter with LocationSIDs that are associated with the stop codes for cardiology (303), renal (313), primary care/internal medicine/hypertension/clinical pharmacist (160, 170, 172, 176, 210, 301, 309, 318, 322, 323, 342, 348, 350, 348, 394), endocrinology/diabetes (304, 306), or cancer (316, 488). If there are multiple readings on one date, take a mean of the readings. SBP values will be dropped if any of the following was true: missing value (either SBP or DBP), systolic less than diastolic, systolic >300 mm Hg, or systolic <60 mm Hg. |
| Diastolic blood pressure_pre6 months | Average of all DBP values in the 6 months prior to the index date (including the index date) corresponding to an outpatient encounter with LocationSIDs that are associated with the stop codes for cardiology (303), renal (313), primary care/internal medicine/hypertension/clinical pharmacist (160, 170, 172, 176, 210, 301, 309, 318, 322, 323, 342, 348, 350, 348, 394), endocrinology/diabetes (304, 306), or cancer (316, 488). If there are multiple readings on one date, take a mean of the readings. SBP values will be dropped if any of the following was true: missing value (either SBP or DBP), systolic less than diastolic, systolic >300 mm Hg, or systolic <60 mm Hg. |
| Heart rate | Most recent proximal heart rate/pulse value corresponding to an outpatient encounter in the one-year pre-index period (including the index date). Heart rate will be set to missing if value is less than 20 or greater than 200. |
| Total cholesterol | The total cholesterol value measured in the outpatient setting closest to the index date in the **five-year** pre-index period (extended from one year due to infrequency of measurement in clinical practice). Defined using OMOPs mapping where LOINC_Mapped is 2093-3 and Topography is (SERUM, PLASMA, BLOOD, SER/PLA, BLOOD*, SER/PLAS, BLOOD., WS-PLASMA, CC SERUM,HIBBING SERUM, MOFH SERUM, OPCC-SERUM,BLOOD VENOUS, LC-SER, SERUM (QUEST)). Values ≥75 mg/dL and ≤500 mg/dL will be retained; outliers will be coded as missing. |
| HDL-C level | The HDL-C value measured in the outpatient setting closest to the index date in the **five-year** pre-index period (extended from one year due to infrequency of measurement in clinical practice). Defined using OMOPs mapping where LOINC_Mapped is 2085-9 and Topography is (SERUM, PLASMA, BLOOD, SER/PLA, BLOOD*, SER/PLAS, BLOOD., WS-PLASMA, CC SERUM,HIBBING SERUM, MOFH SERUM, OPCC-SERUM,BLOOD VENOUS, LC-SER, SERUM (QUEST)). Values ≥10mg/dL and ≤200 mg/dL will be retained; outliers will be coded as missing. |
| LDL-C level | The LDL-C value measured in the outpatient setting closest to the index date in the **five-year** pre-index period (extended from one year due to infrequency of measurement in clinical practice). Defined using OMOPs mapping where LOINC_Mapped is (13457-7,18262-6,2089-1,2574-2,9346-8) and Topography is (SERUM, PLASMA, BLOOD, SER/PLA, BLOOD*, SER/PLAS, BLOOD., WS-PLASMA, CC SERUM,HIBBING SERUM, MOFH SERUM, OPCC-SERUM,BLOOD VENOUS, LC-SER, SERUM (QUEST)). Values ≥25 mg/dL and ≤250 mg/dL will be retained; outliers will be coded as missing. |
| Triglyceride level | The triglyceride value measured in the outpatient setting closest to the index date in the **five-year** pre-index period (extended from one year due to infrequency of measurement in clinical practice). Defined using OMOPs mapping where LOINC_Mapped is 2571-8 and Topography is (SERUM, PLASMA, BLOOD, SER/PLA, BLOOD*, SER/PLAS, BLOOD., WS-PLASMA, CC SERUM,HIBBING SERUM, MOFH SERUM, OPCC-SERUM,BLOOD VENOUS, LC-SER, SERUM (QUEST)). Values ≥20mg/dL and ≤2000 mg/dL will be retained; outliers will be coded as missing. |
| Hemoglobin A1c | The glycated hemoglobin (i.e., “hemoglobin A1c”) value measured in the outpatient setting closest to the index date in the **five-year** pre-index period (extended from one year due to infrequency of measurement in clinical practice). Defined using OMOPs mapping where LOINC_Mapped is 4548-4 and Topography is (SERUM, PLASMA, BLOOD, SER/PLA, BLOOD*, SER/PLAS, BLOOD., WS-PLASMA, CC SERUM,HIBBING SERUM, MOFH SERUM, OPCC-SERUM,BLOOD VENOUS, LC-SER, SERUM (QUEST)). Values ≥3% and ≤20% will be retained; outliers will be coded as missing. |
| Serum creatinine | The serum creatinine value measured in the outpatient setting closest to the index date in the one-year pre-index period. Defined using OMOPs mapping where LOINC_mapped is (2160-0, 38483-4, 77140-2, 40248-7, 44784-7) and Topography in (SERUM, PLASMA, BLOOD, SER/PLA, BLOOD*, SER/PLAS, BLOOD., WS-PLASMA, CC SERUM,HIBBING SERUM, MOFH SERUM, OPCC-SERUM,BLOOD VENOUS, LC-SER, SERUM (QUEST)). Values ≥0.2mg/dL and ≤30 mg/dL will be retained; outliers will be coded as missing. |
| Estimated glomerular filtration rate | The estimated glomerular filtration rate closest to the index date in the one-year pre-index period. This calculation is based on the CKD-EPI 2021 equation that does not include a race coefficient.^6,7^ This calculation can be done manually from age, sex, and serum creatinine; we will also pull the variable eGFR_CKD from the CDW. Values ≥1 mL/min/1.73m^2^ and ≤200 mL/min/1.73m^2^ will be retained; outliers will be coded as missing. |
| Albumin to creatinine ratio | The urine albumin-to-creatinine ratio value measured in the outpatient setting closest to the index date in the **five-year** pre-index period (extended from one year due to infrequency of measurement in clinical practice). Defined using OMOPs mapping where LOINC_Mapped is (30001-2, 32294-1, 77254-1, 34535-5, 9318-7, 2161-8, 14957-5) and Topography is (1 HR URINE, 24 HOUR URINE, 24 HR URINE, 24 HR. URINE, 24-HOUR URINE, 24-HR URINE, 24H URINE, 24H URINE (WR), 24hr URINE, URINE, URINE (24 HOUR), URINE (24 HR), URINE (24HR), URINE (24HRS), URINE (CLEAN CATCH), URINE (RANDOM CHEM), URINE (RANDOM), URINE (SEND-TIMED), URINE (Spot), URINE (STERILE CONTAINER), URINE (T-7X100), URINE [SL], URINE 12 HOUR, URINE 2, URINE 24 HOUR, URINE 24 HR, URINE 24H, URINE 24HR, URINE CLEAN CATCH, URINE RAN, URINE RANDOM, URINE TIMED, URINE(24-HRS), URINE(DRUG SCRN), URINE(DSC), URINE(MICRO, URINE+SERUM, URINE, 24 HOUR, URINE, 24 HR., URINE, 24hr, URINE, CATHETERIZED, URINE, CLEAN CATCH, URINE, RANDOM (SPOT), URINE, SPOT, URINE, TIMED, URINE,24 HOUR, URINE,24 HR., URINE,24HR, URINE,2HR, URINE,C&S, URINE,CATHETER,NEWLY PLACED, URINE,CLEAN CATCH, URINE,RANDOM, URINE,SECOND MORNING COLLECTION, URINE,SPOT, URINE,TIMED, URINE-24, URINE-24 HR, URINE-24hr, URINE-3, URINE-DAU, URINE/24 HR, URINE/24HR [SL], URINE/SERUM, URINE: TIMED). Values ≥0 mg/g and ≤30,000 mg/g will be retained; outliers will be coded as missing. |
| Serum sodium | The serum sodium value measured in the outpatient setting closest to the index date in the one-year pre-index period. Defined using OMOPs mapping where LOINC_Mapped is (2951-2, 34548-8) and Topography is (SERUM, PLASMA, BLOOD, SER/PLA, BLOOD*, SER/PLAS, BLOOD., WS-PLASMA, CC SERUM,HIBBING SERUM, MOFH SERUM, OPCC-SERUM,BLOOD VENOUS, LC-SER, SERUM (QUEST)). Values ≥120 mEq/L and ≤150 mEq/L will be retained; outliers will be coded as missing. |
| Serum potassium | The serum potassium value measured in the outpatient setting closest to the index date in the one-year pre-index period. Defined using OMOPs mapping where LOINC_Mapped is (6298-4, 2823-3, 32713-0) and Topography is (SERUM, PLASMA, BLOOD, SER/PLA, BLOOD*, SER/PLAS, BLOOD., WS-PLASMA, CC SERUM,HIBBING SERUM, MOFH SERUM, OPCC-SERUM,BLOOD VENOUS, LC-SER, SERUM (QUEST)). Values ≥1.5 mEq/L and ≤8 mEq/L will be retained; outliers will be coded as missing. |
| Ejection fraction | Left ventricular ejection fraction (LVEF) value in the **five-year** pre-index period (extended from one year due to infrequency of measurement in clinical practice). If there are multiple measurements on one date or if a range is provided, we will take the average of the two measurements. Values ≥0% and ≤100% will be retained; outliers will be coded as missing. |
| *Comorbidities* |  |
| Alcohol abuse | Either two outpatient claims (7 days apart), or one inpatient claim with at least one of the following diagnosis codes in any position using all available claims prior to the index date (including the index date):   1) ICD-9 codes: 291, 291.0, 291.1, 291.2, 291.4, 291.5, 291.8, 291.81, 291.82, 291.89, 291.9, 303.00, 303.01, 303.02, 303.03, 303.90, 303.91, 303.92, 303.93, 305.00, 305.01, 305.02, 305.03 2) ICD-10 codes: F10.10, F10.120, F10.121, F10.129, F10.14, F10.150, F10.159, F10.180, F10.181, F10.182, F10.188, F10.19, F10.20, F10.21, F10.220, F10.221, F10.229, F10.230, F10.231, F10.232, F10.239, F10.24, F10.250, F10.259, F10.26, F10.27, F10.280, F10.281, F10.282, F10.288, F10.29, F10.920, F10.921, F10.929, F10.94, F10.950, F10.959, F10.96, F10.97, F10.980, F10.981, F10.982, F10.988, F10.99 |
| Angina | Either two outpatient claims (7 days apart), or one inpatient claim with at least one of the following diagnosis codes in any position using all available claims prior to the index date (including the index date):   1) ICD-9 codes: 411.0, 411.1, 411.8, 413.0, 413.1, 413.9 2) ICD-10 codes: I20.0 I20.1, I20.8, I20.9, I25.110, I25.111, I25.118, I25.119, I25.700, I25.701, I25.708, I25.709, I25.710, I25.711, I25.718, I25.719, I25.720, I25.721, I25.728, I25.729, I25.730, I25.731, I25.738, I25.739, I25.750, I25.751, I25.758, I25.759, I25.760, I25.761, I25.768, I25.769, I25.790, I25.791, I25.798, I25.799 |
| Arrhythmia | Either two outpatient claims (7 days apart), or one inpatient claim with at least one of the following diagnosis codes in any position using all available claims prior to the index date (including the index date):   1) ICD-9 codes: 427, 427.0, 427.1. 427.2, 427.3, 427.31, 427.32, 427.4, 427.41, 427.42, 427.5, 427.6, 427.60, 427.61, 427.69, 427.8, 427.81, 427.89, 427.9 2) ICD-10 codes: I47.0, I47.1, I47.2, I47.9, I48.0, I48.1, I48.2, I48.3, I48.4, I48.91, I48.92, I49.01, I49.02, I49.1, I49.2, I49.3, I49.40, I49.49, I49.5, I49.8, I49.9, R00.1 |
| Cancer | Either two outpatient claims (7 days apart), or one inpatient claim with at least one of the diagnosis codes in the following link in any position using all available claims prior to the index date (including the index date): <https://seer.cancer.gov/siterecode/icdo3_dwhoheme/index.html>.^10^ The NCI SEER program captures these ICD-O-3 codes in their registry. Conversion from ICD-O-3 codes to ICD-9 and ICD-10 codes can be found at <https://seer.cancer.gov/tools/conversion/>.^11^  Note: We are requesting access to the VA’s cancer registry. We will evaluate whether cancer should be defined using the registry. The VA uses the software OncoTraX to abstract charts of patients afflicted of cancer to include in the registry back to 1995.^12^ |
| Cardiovascular Disease (CVD) | Meeting criteria for history of heart failure (any definition below), MI, coronary revascularization, peripheral artery disease, or stroke using all available data prior to the index date (including the index date) (see definitions for each below). |
| Chronic kidney disease^13,14^ | Meeting at least one of the following criteria:   1) Either two outpatient claims (7 days apart), or one inpatient claim with at least one of the following diagnosis codes in any position using all available claims prior to the index date (including the index date):    a) ICD-9 codes: 016.0, 095.4, 189.0, 189.9, 223.0, 236.91, 250.4, 271.4, 274.1, 283.11, 403.x1, 403.x0, 404.x2, 404.x3, 404.x0, 404.x1, 440.1, 442.1, 447.3, 572.4, 580–588, 591, 642.1, 646.2, 753.12–753.17, 753.19, 753.2, 794.4    b) ICD-10 codes: A18.11, A52.75, C64.9, C68.9, D30.00, D41.00, D41.20, D59.3, E10.21, E10.29, E11.21, E11.29, E7.48, I120, I12.9, I13.0, I131.0, I131.1, I132, I70.1, I72.2, K76.7, M10.30, N0.03, N0.08, N0.09, N0.13, N0.22, N0.32, N0.33, N0.35, N0.38, N0.39, N0.40, N0.43, N0.44, N0.48, N0.49, N0.52, N0.55, N0.58, N0.59, N08, N13.30, N17.0, N17.1, N17.2, N17.8, N17.9, N18.1, N18.2, N18.3, N18.4, N18.5, N18.6, N18.9, N19, N25.0, N25.1, N25.81, N25.89, N25.9, N26.9,Q61.02, Q6119, Q61.2, Q61.3, Q61.4, Q61.5, Q61.8, Q621.0, Q621.1, Q621.2, Q62.31, Q62.39, R94.4. 2) Estimated glomerular filtration rate of <60 mL/min/1.73 m^2^ on 2 separate visits at least 30 days or more apart in the one-year pre-index period. To account for data/reading errors, only eGFR values between 0 and 250 will be considered. 3) Meets criteria for end-stage kidney disease or dialysis, as below. |
| Chronic Liver Disease | Either two outpatient claims (7 days apart), or one inpatient claim with at least one of the following diagnosis codes in any position using all available claims prior to the index date (including the index date):   1) ICD-9 codes: 070.22, 070.23, 070.32, 070.33, 070.44, 070.54, 456.0, 456.1, 456.20, 456.21, 570.x, 571.x, 572.x, 573.x, V42.7 2) ICD-10 codes: B18.0-B18.2, B25.1, I85.00, I85.01, I85.10, I85.11, K70.0, K70.10, K70.11, K70.2, K70.30, K70.31, K70.40, K70.41, K70.9, K71.0, K71.10, K71.11, K71.2, K71.3, K71.50, K71.51, K71.6, K71.7, K71.8, K71.9, K72.00, K72.01, K72.10, K72.11, K72.90, K72.91, K73.0, K73.1, K73.2, K73.8, K73.9, K74.0, K74.1, K74.2, K74.3, K74.4, K74.5, K74.60, K74.69, K75.0, K75.1m K75.2, K75.3, K75.4, K75.81, K75.89, K75.9, K76.0, K76.1, K76.2, K76.3, K76.4, K76.5, K76.6, K76.7, K76.81, K76.89, K76.9, K77.x, Z48.23, Z94.4 |
| Chronic Lung Disease | Either two outpatient claims (7 days apart), or one inpatient claim with at least one of the following diagnosis codes in any position using all available claims prior to the index date (including the index date):   1) ICD-9 codes: 490, 491, 491.0, 491.1, 491.2, 491.20, 491.21, 491.22, 491.8, 491.9, 492, 492.0, 492.8, 493, 493.0, 493.00, 493.01, 493.02, 493.1, 493.10, 493.11, 493.12, 493.2, 493.20, 493.21, 493.22, 493.9, 493.90, 493.91, 493.92 494.0, 494.1, 495.0, 495.1, 495.2, 495.3, 495.4, 495.5, 495.6, 495.7, 495.8, 495.9, 496, 500, 501, 502, 503, 504, 505, 506.4 2) ICD-10 codes: J40, J41.0, J41.1, J41.8, J42, J43.0, J43.1, J43.2, J43.8, J43.9, J44.0, J44.1, J44.9, J45.20, J45.21, J45.22, J45.30, J45.31, J45.32, J45.40, 45.41, J45.42, J45.50, J45.51, J45.52, J45.901, J45.902, J45.909, J45.998, J47.0, J47.1, J47.9, J60, J61, J62.0, J62.8, J63.0, J63.1, J63.2, J63.3, J63.4, J63.5, J63.6, J64, J65, J66.0, J66.1, J66.2, J66.8, J67.0, J67.1, J67.2, J67.3, J67.4, J67.5, J67.6, J67.7, J67.8, J67.9, J68.4 |
| Cirrhosis | Either two outpatient claims (7 days apart), or one inpatient claim with at least one of the following diagnosis codes in any position using all available claims prior to the index date (including the index date):   1) ICD-9 codes: 070.0, 070.2, 070.20, 070.21, 070.4, 070.41, 070.42, 070.43, 070.49, 070.6, 070.71, 571.2, 571.5, 571.6 2) ICD-10 codes: B15.0, B16.0, B16.2, B17.11, B19.0, B19.11, B19.21 K70.30, K70.31, K70.41, K71.11, K71.7, K72.01, K72.11, K72.91, K74.3, K74.4, K74.5, K74.60, K74.69, P78.81 |
| Prior coronary revascularization | Meeting BOTH criteria 1 and 2 below.   1) Defined by ≥1 inpatient or outpatient procedure with one of the following using all available claims prior to the index date (including the index date):    a) CPT code for coronary revascularization: 33510-33519, 33521-33523, 33530, 33533-33536, 92920, 92921, 92924, 92925, 92928, 92929, 92933, 92934, 92937, 92938, 92941, 92943, 92944, 92980, 92981, 92982, 92984, or 92996    b) ICD-9 procedure code (any position) of 00.66, 36.0, 36.10-36.19, or 36.2    c) ICD-10 procedure code starting with any of the following: 0210, 0211, 0212, 0213, 0270, 0271, 0272, 0273, 02C0, 02C1, 02C2, 02C3, or 3E07. 2) Meet at least one of the following criteria:    a) Have no inpatient claims with a discharge diagnosis code for acute myocardial infarction (ICD-9 codes 410.x0 or 410.x1 or ICD-10 codes I21.xx or I22.xx) within 60 days prior to the procedure.    b) Have primary discharge diagnosis codes for non-elective CHD-related hospitalization prior to the index date (including the index date): a. Arrhythmia: ICD-9 diagnosis code of 427.xx [except 427.5] or ICD-10 diagnosis code of I47.1, I47.2, I47.9, I48.91, I48.92, I49.01, I49.02, I49.1, I49.3, I49.40, I49.49, I49.5, I49.8, I49.9, R00.1. b. Cardiac arrest: ICD-9 diagnosis code of 427.5, or ICD-10 diagnosis code of I46.9. c. Heart failure: ICD-9 diagnosis code of 402.01, 402.11, 402.91, 404.01, 404.03, 404.11, 404.13, 404.91, 404.93, or 428.x, or ICD-10 diagnosis code of I11.0, I13.0, I13.2, I50.1, I50.20, I50.21, I50.22, I50.23, I50.30, I50.31, I50.32, I50.33, I50.40, I50.41, I50.42, I50.43, or I50.9. d. Unstable angina: ICD-9 diagnosis code of 411.xx or ICD-10 diagnosis code of I20.0, I24.0, I24.1, I24.8. |
| Diabetes | Meeting one of the two criteria below:   1) Either two outpatient claims (7 days apart), or one inpatient claim with at least one of the following diagnosis codes in any position using all available claims prior to the index date (including the index date):    a) ICD-9 codes: 250.xx, 357.2, 362.0x, or 366.41.    b) ICD-10 codes: E08.36, E08.42, E09.36, E09.42, E10.10, E10.11, E10.29, E10.311, E10.319, E10.36, E10.39, E10.40, E10.42, E10.51, E10.618, E10.620, E10.621, E10.622, E10.628, E10.630, E10.638, E10.641, E10.649, E10.65, E10.69, E10.8, E10.9, E11.00, E11.01, E11.29, E11.311, E11.319, E11.329, E11.339, E11.349, E11.359, E11.36, E11.39, E11.40, E11.42, E11.51, E11.618, E11.620, E11.621, E11.622, E11.628, E11.630, E11.638, E11.641, E11.649, E11.65, E11.69, E11.8, E11.9, E13.10, E13.36, E13.42. 2) ≥1 pharmacy claim for an oral antidiabetic drug fill or insulin in the one-year prior to the index date (including the index date). |
| Depression^15^ | Either two outpatient claims (7 days apart), or one inpatient claim with at least one of the following diagnosis codes in any position using all available claims prior to the index date (including the index date):   1) ICD-9 codes: 296.2, 296.3, 296.5, 300.4, 309.x, or 311. 2) ICD-10 codes: F20.4, F31.3-F31.5, F32.x, F33.x, F34.1, F41.2, or F43.2. |
| Drug/substance abuse | Either two outpatient claims (7 days apart), or one inpatient claim with at least one of the following diagnosis codes in any position using all available claims prior to the index date (including the index date):   1) ICD-9 codes: 292.0, 292.8x, 292.9, 304.0x, 304.1x, 304.2x, 304.3x, 304.4x, 304.5x, 304.6x, 304.7x, 304.8x, 304.9x, 305.2x, 305.3x, 305.4x, 305.5x, 305.6x, 305.7x, 305.8x, 305.9x, 648.3x 2) ICD-10 codes: F11.x, F12.x, F13.x, F14.x, F15.x, F16.x, F17.x, F18.x, F19.x, F55.x, O99.32x |
| Frailty status^16^ | Defined using the 31-item VA Frailty Index (VA-FI) (see below). Based on the VA-FI, patients will be categorized into non-frail (FI ≤0.21) or frail (FI >0.21). |
| VA Frailty Index (VA-FI) | 31-item index used to define frailty. Ratio of the sum of the number of health deficits to the total number of health factors evaluated.^17-24^ For example, a Veteran with 10 of 30 possible deficits would have an FI of 10/30 = 0.33. The 31 variables used to calculate the VA-FI have been validated in the Veterans Health Affairs system as described by Orkaby et al.^24^ The variables and codes used to calculate the VA-FI are shown in Appendix F. |
| Heart failure with reduced ejection fraction (systolic HF) | Meeting one of the three criteria below. Criteria below are sequentially applied.   1) PREFERENCE: Left ventricular ejection fraction (LVEF) value ≤40 in the **five-year** pre-index period (extended from one year due to infrequency of measurement in clinical practice). If there are multiple measurements on one date or if a range is provided, we will take the average of the two measurements. 2) If no LVEF measurement is available, then we will look for either two outpatient claims (7 days apart), or one inpatient claim with at least one of the following diagnosis codes in any position using all available claims prior to the index date (including the index date):    a) ICD-9 codes: 428.0x, 428.1x, 428.2x, or 428.4x.    b) ICD-10 codes: I50.1, I50.2x, I50.4x, or I50.9. 3) Finally, if a patient does not have an LVEF or one of the codes above, we will look for at least one prescription for sacubitril/valsartan in the 104 days (90 days + 14-day grace period) prior to the index date (including the index date). |
| Heart failure with preserved ejection fraction (diastolic HF) | Meeting both criteria below.   1) Left ventricular ejection fraction (LVEF) value >40 in the **five-year** pre-index period (extended from one year due to infrequency of measurement in clinical practice). If there are multiple measurements on one date or if a range is provided, we will take the average of the two measurements. 2) Two outpatient claims (at least 7 days apart), or one inpatient claim with at least one of the following diagnosis codes in any position using all available claims prior to the index date (including the index date):    a) ICD-9 code: 428.3x.    b) ICD-10 codes: I50.3. |
| Ischemic heart failure | Either two outpatient claims (7 days apart), or one inpatient claim with at least one of the following diagnosis codes in any position using all available claims prior to the index date (including the index date):   1) ICD-9 codes: 428.xx, 414.8 2) ICD-10 codes: I50.XXX, I25.5 |
| Non-ischemic heart failure | Either two outpatient claims (7 days apart), or one inpatient claim with at least one of the following diagnosis codes in any position using all available claims prior to the index date (including the index date):   1) ICD-9 codes: 398.91, 402.X1, 404.X1, 404.X3, 415, 416.9, 422, 425.xx, 425.8 2) ICD-10 codes: I09.81, I11.0, I13.0, I13.2, I26.0X, I27.81, I27.9, I41, I42.X, I43.0 |
| Myocardial infarction | Either two outpatient claims (7 days apart), or one inpatient claim with at least one of the following diagnosis codes in any position using all available claims prior to the index date (including the index date):   1) ICD-9 codes: 410.x 2) ICD-10 codes: I21.x, I22.x, or I25.2 |
| Obstructive sleep apnea | Either two outpatient claims (7 days apart), or one inpatient claim with at least one of the following diagnosis codes in any position using all available claims prior to the index date (including the index date):   1) ICD-9 code: 327.2X, 327.23, 327.29, 780.51, 780.53, 780.57 2) ICD-10 code: G47.30, G47.33, G47.39 |
| Peripheral Arterial Disease (PAD) | Either two outpatient claims (7 days apart), or one inpatient claim with at least one of the following diagnosis codes in any position using all available claims prior to the index date (including the index date) Meeting one of the two criteria below:   1) ICD codes:    a) ICD-9 codes: 440.20-440.24, 440.31, 444.2, 443.9, or 444.81.    b) ICD-10 codes: I70.209, I70.219, I70.229, I70.25, I70.269, I70.499, I73.9.    c) ICD-9 procedure codes: 38.x, 39.22, 39.24, 29.36, 39.26, 39.28, 39.5, or 39.50. 2) CPT codes: 37205 or 75962. |
| Any Stroke^2^ | Meeting one of the two criteria below using all available claims prior to the index date (including the index date):   1) ICD-9 codes:    a) ≥1 inpatient claim with a discharge diagnosis code (primary or secondary position) of 431, 432.X, 433.x1, 434.xx, 433.x0, 436, 437.x, 438.xx.    b) ≥1 outpatient claim with a diagnosis code (any position) of 431, 432.X, 433.x1, 434.xx, 433.x0, 436, 437.x, 438.xx. 2) ICD-10 codes:    a) ≥1 inpatient discharge diagnosis code (primary or secondary position) of I60.xx, I61.x, I62.xx, I63.xxx, I66.xx, I65.xx, I67.xxx, I68.x, I69.xxx    b) ≥1 outpatient claim with diagnosis code (any position) of I60.xx, I61.x, I62.xx, I63.xxx, I66.xx, I65.xx, I67.xxx, I68.x, I69.xxx    c) ≥1 inpatient ICD-10 procedure code of 03CH0ZZ, 03CH4ZZ, 03CJ0ZZ, 03CJ4ZZ, 03CK0ZZ, 03CK4ZZ, 03CL0ZZ, 03CL4ZZ, 03CM0ZZ, 03CM4ZZ, 03CN0ZZ, 03CN4ZZ, 03RH07Z, 03RH0JZ, 03RH0KZ, 03RH47Z, 03RH4JZ, 03RH4KZ, 03RJ07Z, 03RJ0JZ, 03RJ0KZ, 03RJ47Z, 03RJ4JZ, 03RJ4KZ,03RK07Z, 03RK0JZ, 03RK0KZ, 03RK47Z, 03RK4JZ, 03RK4KZ, 03RL07Z, 03RL0JZ, 03RL0KZ, 03RL47Z, 03RL4JZ, 03RL4KZ, 03RM07Z, 03RM0JZ, 03RM0KZ, 03RM47Z, 03RM4JZ, 03RM4KZ, 03RN07Z, 03RN0JZ, 03RN0KZ, 03RN47Z, 03RN4JZ, or 03RN4KZ. 3) CPT codes: ≥1 inpatient or outpatient claim with a CPT code for carotid revascularization of 35301, 35390, 37215, 37216, 0005T, 0075T, or 0076. |
| Ischemic stroke^3,4^ | Meeting one of the two criteria below using all available claims prior to the index date (including the index date):   1) ICD-9 codes:    a) ≥1 inpatient claim with a discharge diagnosis code (primary or secondary position) of 433.x1, 434.xx, 433.x0, 436, 437.x, 438.xx.    b) ≥1 outpatient claim with a diagnosis code (any position) of 433.x1, 434.xx, 433.x0, 436, 437.x, 438.xx. 2) ICD-10 codes:    a) ≥1 inpatient discharge diagnosis code (primary or secondary position) of I63.xxx, I66.xx, I65.xx, I67.xxx, I68.x, I69.3X    b) ≥1 outpatient claim with diagnosis code (any position) of I63.xxx, I66.xx, I65.xx, I67.xxx, I68.x, I69.xxx    c) ≥1 inpatient ICD-10 procedure code of 03CH0ZZ, 03CH4ZZ, 03CJ0ZZ, 03CJ4ZZ, 03CK0ZZ, 03CK4ZZ, 03CL0ZZ, 03CL4ZZ, 03CM0ZZ, 03CM4ZZ, 03CN0ZZ, 03CN4ZZ, 03RH07Z, 03RH0JZ, 03RH0KZ, 03RH47Z, 03RH4JZ, 03RH4KZ, 03RJ07Z, 03RJ0JZ, 03RJ0KZ, 03RJ47Z, 03RJ4JZ, 03RJ4KZ,03RK07Z, 03RK0JZ, 03RK0KZ, 03RK47Z, 03RK4JZ, 03RK4KZ, 03RL07Z, 03RL0JZ, 03RL0KZ, 03RL47Z, 03RL4JZ, 03RL4KZ, 03RM07Z, 03RM0JZ, 03RM0KZ, 03RM47Z, 03RM4JZ, 03RM4KZ, 03RN07Z, 03RN0JZ, 03RN0KZ, 03RN47Z, 03RN4JZ, or 03RN4KZ. 3) CPT codes: ≥1 inpatient or outpatient claim with a CPT code for carotid revascularization of 35301, 35390, 37215, 37216, 0005T, 0075T, or 0076. |
| Hemorrhagic stroke^3,4^ | Meeting one of the two criteria below using all available claims prior to the index date (including the index date):   1) ICD-9 codes:    a) ≥1 inpatient claim with a discharge diagnosis code (primary or secondary position) of 431, or 432.X    b) ≥1 outpatient claim with a diagnosis code (any position) of 431, or 432.X 2) ICD-10 codes:    a) ≥1 inpatient discharge diagnosis code (primary or secondary position) of I60.xx, I61.x, I62.xx, I69.0X, I69.1X, I69.2X    b) ≥1 outpatient claim with diagnosis code (any position) of I60.xx, I61.x, I62.xx, I69.xxx |
| Syncope | Either two outpatient claims (7 days apart), or one inpatient claim with at least one of the following diagnosis codes in any position using all available claims prior to the index date (including the index date):   1) ICD-9 code: 780.2x 2) ICD-10 code: R55.x |
| *Concomitant Medication Use* | *****Please also pull dates for each medication.*** |
| Statin use | Defined as one or more outpatient pharmacy dispenses for a statin medication in the 104 days (90 days + 14-day grace period) prior to each Veteran’s index date (including the index date). Statin medications include: atorvastatin, rosuvastatin, simvastatin, fluvastatin, pitavastatin, pravastatin, or lovastatin. |
| Aspirin use | Defined as one or more outpatient pharmacy dispenses for aspirin in the 104 days (90 days + 14-day grace period) prior to each Veteran’s index date (including the index date). |
| SGLT2 inhibitor use | Defined as one or more outpatient pharmacy dispenses for an SGLT-2 inhibitor medication in the 104 days (90 days + 14-day grace period) prior to each Veteran’s index date (including the index date). SGLT-2 inhibitor medications include: canagliflozin, dapagliflozin, empagliflozin, and ertugliflozin. |
| Antidepressant use | Defined as one or more outpatient pharmacy dispenses for an antidepressant medication in the 104 days (90 days + 14-day grace period) prior to each Veteran’s index date (including the index date). Antidepressant medications include: amitriptyline, amoxapine, bupropion, citalopram, clomipramine, desipramine, desvenlafaxine, escitalopram, fluoxetine, fluoxetine, fluvoxamine, imipramine, isocarboxazid, levomilnacipran, maprotiline, milnacipran, mirtazaine, moclobemide, nefazodone, nortriptyline, paroxetine, phenelzine, protriptyline, selegiline, sertraline, tranylcypromine, trazodone, trimipramine, venlafaxine, vilazodone, vortioxetine. |
| Non-ACEI or ARB antihypertensive class | Defined as one or more outpatient pharmacy dispense for an oral non-ACEI or ARB medication class in the pre-index period with a days’ supply that overlaps with the Veteran’s index date (including the index date). Categorized by class as aldosterone receptor antagonist, beta-blocker, calcium channel blocker, centrally-acting drug, direct arterial vasodilator, direct renin inhibitor, thiazide diuretic, loop diuretic, and potassium sparing diuretic. See Appendix E for specific drug names within each class. Exclude all non-oral products, except for clonidine patches. |
| Number of antihypertensive medications | Sum of the number of antihypertensive medications dispensed in the pre-index period with a days’ supply that overlaps with the Veteran’s index date (including the index date). |
| *Healthcare utilization* |  |
| Number of primary care visits | Defined as the continuous number of outpatient primary care encounters (160, 170, 172, 176, 210, 301, 309, 318, 322, 323, 342, 348, 350, 348, 394) in the one-year pre-index period (including the index date). |
| Number of hospitalizations | Defined as the continuous number of all-cause hospitalizations in the one-year pre-index period (including the index date). |
| Number of emergency department visits | Defined as the continuous number of emergency department visits in the one-year pre-index period (including the index date). |
| *Facility-level Covariates* |  |
| Primary center | Sta3n for the Veteran. |
| Academic setting | Coded for each sta3n as academic or non-academic based on <http://www.friendsofva.org/resources/2012/finalvainfrastructurereport.pdf>. |
| Urban setting | Coded for each sta3n as rural or non-rural based on <https://www.ruralhealth.va.gov/docs/atlas/CHAPTER_02_RHRI_Pts_treated_at_VAMCs.pdf>. Facilities with red lettering in Table 2 are coded as rural. |
| Volume of hypertensive patients at center | Number of incident hypertensive patients for each Sta3n for the Veteran. |

#### eTable 2. Definitions of Dementia Outcomes. Criteria for identifying dementia outcomes from ICD codes and medication records.

| **Variable** | **Definition** |
| --- | --- |
| *Primary outcomes* |  |
| Alzheimers Disease and Related Dementias (ADRD)^8,9^ | Meet one of the three of the criteria below using all available claims. For ICD codes, patients had to have either two outpatient claims (7 days apart), or one inpatient claim with at least one of the following codes in any position.   1) ICD codes:    a) ICD-9 codes: 290.0, 290.1x, 290.2x, 290.3, 90.4x, 290.8, 290.9, 291.1, 291.2, 292.82, 294.0, 294.10, 294.11, 294.20, 331.0, 331.82, 331.11, 331.19, 333.0, 797    b) ICD-10 codes: F06.1, F06.8, F07.0, G13.2, G13.8, G23.0, G23.1, G23.2, G23.8, G23.9, G30.8, G30.9, G31.01, G31.09, G31.1, G31.2, G31.83, G31.85, G31.89, G31.9, G94, R41.81, R54 2) Dispense of at least one dementia medication: memantine, galantamine, rivastigmine, or donepezil. 3) Natural Language Processing (limited set per Reuben et al^8^ – expansion to occur at later date)    a) presence of the terms “dementia” or “neurodegenerative” without the presence of any of the following markers in the same statement:       i) Negating words, such as “not”, “negative”, or “ruled out”       ii) Words that referred to the patient’s family history or a family member’s dementia status, such as “family history”, “wife”, or “husband”       iii) Words that indicated uncertainty, such as “suspected”, “possible”, or “risk” |

#### eTable 3. Variables used to define study population

| **Variable** | **Definition** |
| --- | --- |
| Index date | Date of the first-ever outpatient pharmacy dispense of either an angiotensin-II receptor blocker (ARB)- or an angiotensin-converting enzyme inhibitor (ACEI) (i.e., new users) in the index date identification period. |
| Index date identification period | January 1, 2000, to December 31, 2017 (dates inclusive). |
| Index exposure group: ARB | Angiotensin-II receptor blocker. Patients are categorized into this exposure group irrespective of other non-ACEI or ARB medications dispensed. |
| Index exposure group: ACEI | Angiotensin converting enzyme inhibitor. Patients are categorized into this exposure group irrespective of other non-ACEI or ARB medications dispensed. The ACEI group serves as the referent group. |
| Index exposure medication name | Name of medication dispensed on index date. |
| Index exposure dose | Dose of medication dispensed on index date, described as milligrams of drug taken per day. We will first parse the dosage value from LocalDrugNameWithDose then use the QtyNumeric and DaysSupply variables to inform the parsed dose value (if partial or multiple pills are being taken per day, for example). |
| Hypertension | Any of the following using all available outpatient claims in the one year prior to, or 180 days after, the index date, including the index date). *Please note: only 1 of the 2 diagnoses needs to occur in the one year prior to the index date.*   1) ICD-9 codes:    a) ≥1 outpatient claims with a diagnosis code (any position) of 401.x, 403.0x, 403.1x, 403.9x. 2) ICD-10 codes:    a) ≥1 outpatient claims with a diagnosis code (any position) of I10, I12.0, I12.9. |

#### eTable 4. Baseline Characteristics of Veterans Before and After Inverse Probability Weighting. Continuous variables are summarized as mean (SD) or median (IQR), and categorical variables as counts (percentages). Absolute standardized mean differences (ASMD) <0.10 indicate acceptable balance.

|  | **Before IP Weighting** | | | **After IP Weighting** | | |
| --- | --- | --- | --- | --- | --- | --- |
| **Variable** | **ARB**  N = 247,802^1^ | **ACEI**  N = 2,329,198^1^ | **ASMD** | **ARB** | **ACEI** | **ASMD** |
| **Treatment initiation (years)** | 2,010 (2,004, 2,014) | 2,007 (2,004, 2,011) | 0.32 | 2007 (2003, 2012) | 2007 (2004, 2012) | 0.02 |
| **Age (years)** | 65 (12) | 63 (12) | 0.20 | 64 (12) | 63 (12) | 0.08 |
| **Male sex** |  |  | 0.05 |  |  | 0.01 |
| F | 13,616 (5.5%) | 101,354 (4.4%) |  | 10393.9 (4.6%) | 103065.8 (4.4%) |  |
| M | 234,186 (95%) | 2,227,844 (96%) |  | 215529.8 (95%) | 2213566.1 (96%) |  |
| **Race/Ethnicity** |  |  | 0.06 |  |  | 0.048 |
| Hispanic | 8,973 (3.6%) | 107,160 (4.6%) |  | 8979.4 (4%) | 104558 (4.5%) |  |
| Non-Hispanic Black | 35,398 (14%) | 352,508 (15%) |  | 32095.4 (14%) | 348626.6 (15%) |  |
| Non-Hispanic White | 162,101 (65%) | 1,501,069 (64%) |  | 146198.6 (65%) | 1494292.6 (65%) |  |
| Other | 41,330 (17%) | 368,461 (16%) |  | 38650.1 (17%) | 369154.7 (16%) |  |
| **County-level income** | 37,442 (30,925, 45,026) | 34,761 (29,180, 41,596) | 0.24 | 34828.5 (28959, 42219) | 34753.5 (29118, 41689.5) | 0.02 |
| **Veterans integrated service network region** |  |  | 0.17 |  |  | 0.03 |
| Northeast | 49,109 (20%) | 462,373 (20%) |  | 46635.9 (21%) | 461778.4 (20%) |  |
| Continental | 75,527 (30%) | 648,557 (28%) |  | 65172 (29%) | 650566.3 (28%) |  |
| Southeast | 72,749 (29%) | 848,244 (36%) |  | 77578.8 (34%) | 831152.4 (36%) |  |
| Pacific | 50,417 (20%) | 370,024 (16%) |  | 36536.8 (16%) | 373134.8 (16%) |  |
| **Supplemental insurance type** |  |  | 0.33 |  |  | 0.051 |
| MEDICAIDE | 363 (0.1%) | 2,452 (0.1%) |  | 297.2 (0.1%) | 2554.2 (0.1%) |  |
| MEDICARE | 20,483 (8.3%) | 81,075 (3.5%) |  | 10913.7 (4.8%) | 91619.9 (4%) |  |
| None | 19,582 (7.9%) | 98,908 (4.2%) |  | 11931.1 (5.3%) | 106671.8 (4.6%) |  |
| PRIVATE | 11,961 (4.8%) | 43,341 (1.9%) |  | 5654.3 (2.5%) | 49542.3 (2.1%) |  |
| Unknown | 195,413 (79%) | 2,103,422 (90%) |  | 197127.4 (87%) | 2066243.7 (89%) |  |
| **Priority group status** |  |  | 0.05 |  |  | 0.01 |
| 1(highest need) | 37,890 (17%) | 324,125 (15%) |  | 35499 (16%) | 355105.1 (15%) |  |
| 2 through 9 | 183,369 (83%) | 1,791,567 (85%) |  | 190424.5 (84%) | 1961526.6 (85%) |  |
| **Current smoker** | 28,697 (12%) | 444,067 (19%) | 0.21 | 36147.9 (16%) | 426879.6 (18%) | 0.07 |
| **Homeless or history of homelessness** | 3,690 (1.5%) | 64,308 (2.8%) | 0.09 | 4875.3 (2.2%) | 61512.8 (2.7%) | 0.03 |
| **Body mass index, kg/m2** | 31.2 (5.9) | 30.5 (6.0) | 0.12 | 30.7 (5.8) | 30.5 (6) | 0.03 |
| **Systolic blood pressure, mm Hg** | 141 (18) | 144 (18) | 0.14 | 143 (18) | 144 (18) | 0.02 |
| **Diastolic blood pressure, mm Hg** | 80 (12) | 82 (12) | 0.20 | 81 (12) | 82 (12) | 0.07 |
| **Heart rate, beats/minute** | 74 (13) | 75 (14) | 0.07 | 75 (14) | 75 (14) | 0.02 |
| **Total cholesterol, mg/dL** | 179 (42) | 186 (43) | 0.17 | 184 (43) | 185 (43) | 0.04 |
| **High-density lipoprotein cholesterol, mg/dL** | 45 (14) | 45 (14) | 0.02 | 45 (14) | 45 (15) | 0 |
| **Low-density lipoprotein cholesterol, mg/dL** | 104 (35) | 110 (35) | 0.17 | 108 (36) | 109 (36) | 0.04 |
| **Triglycerides, mg/dL** | 133 (92, 196) | 136 (93, 204) | 0.06 | 138.5 (96, 199) | 138 (95, 200) | 0.01 |
| **Serum creatinine, mg/dL** | 1.05 (0.90, 1.22) | 1.00 (0.90, 1.20) | 0.14 | 1.02 (0.9, 1.2) | 1 (0.9, 1.2) | 0.06 |
| **Hemoglobin A1c** |  |  | 0.07 |  |  | 0.02 |
| < 5.7% | 27,842 (11%) | 248,465 (11%) |  | 24135.2 (11%) | 248938.7 (11%) |  |
| 5.7% - < 6.5% | 40,987 (17%) | 361,932 (16%) |  | 35536.9 (16%) | 362914.7 (16%) |  |
| 6.5% - < 8.0% | 29,481 (12%) | 266,691 (11%) |  | 26860.4 (12%) | 266875.4 (12%) |  |
| 8.0% - < 10.0% | 10,984 (4.4%) | 107,981 (4.6%) |  | 10570 (4.7%) | 107367.8 (4.6%) |  |
| $\geq$ 10% | 4,310 (1.7%) | 58,878 (2.5%) |  | 4938.6 (2.2%) | 57018.2 (2.5%) |  |
| Unknown | 134,198 (54%) | 1,285,251 (55%) |  | 123882.4 (55%) | 1273517.3 (55%) |  |
| **Estimated glomerular filtration rate, ml/min/1.73m^2** | 78 (63, 93) | 83 (68, 96) | 0.22 | 80 (65, 95) | 81 (67, 96) | 0.09 |
| **Urinary albumin to creatinine ratio** |  |  | 0.03 |  |  | 0.02 |
| < 30 mg/g | 20,009 (8.1%) | 185,475 (8.0%) |  | 17973.9 (8%) | 184906.6 (8%) |  |
| 30 - < 300 mg/g | 27,077 (11%) | 238,765 (10%) |  | 24463 (11%) | 239954.5 (10%) |  |
| 300 - < 1000 mg/g | 1,404 (0.6%) | 12,258 (0.5%) |  | 1223.8 (0.5%) | 12341.8 (0.5%) |  |
| $\geq$ 1000 mg/g | 177 (<0.1%) | 1,076 (<0.1%) |  | 131 (<0.1%) | 1137.9 (<0.1%) |  |
| Unknown | 199,135 (80%) | 1,891,624 (81%) |  | 182131.5 (0.81) | 1878291.2 (0.81) |  |
| **Serum sodium, mEq/L** | 139.3 (2.8) | 139.2 (2.9) | 0.03 | 139.3 (2.9) | 139.2 (2.9) | 0.01 |
| **Serum potassium, mEq/L** | 4.24 (0.45) | 4.24 (0.44) | 0.00 | 4.25 (0.44) | 4.24 (0.44) | 0.01 |
| **Ejection fraction** |  |  | 0.09 |  |  | 0.05 |
| $\leq$ 40% | 8,515 (3.4%) | 90,199 (3.9%) |  | 9139.2 (4%) | 89415.4 (3.9%) |  |
| 40% - 50% | 5,113 (2.1%) | 62,835 (2.7%) |  | 7363.8 (3.3%) | 62496.2 (2.7%) |  |
| > 50% | 22,034 (8.9%) | 254,579 (11%) |  | 26779.5 (12%) | 253182.6 (11%) |  |
| Unknown | 212,140 (86%) | 1,921,585 (82%) |  | 182641.3 (81%) | 1911537.5 (83%) |  |
| **Cardiovascular disease** | 62,336 (25%) | 245,541 (11%) | 0.39 | 26990.8 (12%) | 265521.3 (11%) | 0.01 |
| **Myocardial infarction** | 597 (0.2%) | 16,355 (0.7%) | 0.07 | 1006 (0.4%) | 15385 (0.7%) | 0.03 |
| **Peripheral arterial disease** | 3,106 (1.3%) | 44,637 (1.9%) | 0.05 | 4107.9 (1.8%) | 43402.6 (1.9%) | 0 |
| **Any stroke** | 10,427 (4.2%) | 106,048 (4.6%) | 0.02 | 10396.8 (4.6%) | 105153.8 (4.5%) | 0 |
| **Prior coronary revascularization** | 239 (<0.1%) | 6,855 (0.3%) | 0.04 | 429 (0.2%) | 6437.2 (0.3%) | 0.02 |
| **Any heart failure** | 54,126 (22%) | 135,082 (5.8%) | 0.48 | 17097 (7.6%) | 158168.2 (6.8%) | 0.02 |
| **Alcohol abuse** | 4,415 (1.8%) | 89,984 (3.9%) | 0.13 | 6260.4 (2.8%) | 85364.2 (3.7%) | 0.06 |
| **Angina** | 1,034 (0.4%) | 23,010 (1.0%) | 0.07 | 1702.8 (0.8%) | 21870.7 (0.9%) | 0.02 |
| **Arrhythmia** | 8,071 (3.3%) | 82,748 (3.6%) | 0.02 | 9792.3 (4.3%) | 83372.7 (3.6%) | 0.04 |
| **Cancer** | 1,161 (0.5%) | 17,649 (0.8%) | 0.04 | 1515 (0.7%) | 17045.6 (0.7%) | 0.01 |
| **Chronic kidney disease** | 9,743 (3.9%) | 65,539 (2.8%) | 0.06 | 8544.3 (3.8%) | 68640.8 (3%) | 0.05 |
| **Chronic liver disease** | 2,018 (0.8%) | 32,982 (1.4%) | 0.06 | 2604.6 (1.2%) | 31718.1 (1.4%) | 0.02 |
| **Chronic lung disease** | 12,744 (5.1%) | 158,046 (6.8%) | 0.07 | 15815.4 (7%) | 155538.3 (6.7%) | 0.01 |
| **Cirrhosis** | 399 (0.2%) | 6,475 (0.3%) | 0.02 | 539.1 (0.2%) | 6242.2 (0.3%) | 0.01 |
| **Diabetes** | 37,868 (15%) | 398,971 (17%) | 0.05 | 39746.8 (18%) | 395186.9 (17%) | 0.01 |
| **Depression** | 19,272 (7.8%) | 251,125 (11%) | 0.10 | 22134.3 (9.8%) | 244334.4 (11%) | 0.03 |
| **Drug/substance use** | 2,776 (1.1%) | 56,462 (2.4%) | 0.10 | 3977.4 (1.8%) | 53630.4 (2.3%) | 0.04 |
| **Obstructive sleep apnea** | 8,577 (3.5%) | 68,099 (2.9%) | 0.03 | 7692.9 (3.4%) | 69682.7 (3%) | 0.02 |
| **Syncope** | 488 (0.2%) | 8,143 (0.3%) | 0.03 | 738.7 (0.3%) | 7870.7 (0.3%) | 0 |
| **Statin** | 108,629 (44%) | 992,405 (43%) | 0.02 | 98374.6 (44%) | 990725.1 (43%) | 0.02 |
| **Aspirin** | 27,862 (11%) | 327,480 (14%) | 0.08 | 30581.2 (14%) | 321581.4 (14%) | 0.01 |
| **Sodium-glucose transporter type 2 inhibitor** | 69 (<0.1%) | 173 (<0.1%) | 0.02 | 25.1 (0%) | 213 (0%) | 0 |
| **Antidepressant** | 39,376 (16%) | 440,382 (19%) | 0.08 | 40118.2 (18%) | 432515.1 (19%) | 0.02 |
| **Number of antihypertensive medications** | 2 (1, 3) | 2 (1, 2) | 0.14 | 2 (1, 3) | 2 (1, 2) | 0.07 |
| **Alpha-blocker** | 20,472 (8.3%) | 203,552 (8.7%) | 0.02 | 20846.9 (9.2%) | 201995 (8.7%) | 0.02 |
| **Beta-blocker** | 70,320 (28%) | 651,970 (28%) | 0.01 | 66694.5 (30%) | 651949.7 (28%) | 0.03 |
| **Calcium channel blocker** | 63,062 (25%) | 459,706 (20%) | 0.14 | 50325.1 (22%) | 469458.1 (20%) | 0.05 |
| **Centrally-acting drug** | 5,420 (2.2%) | 36,317 (1.6%) | 0.05 | 4158.5 (1.8%) | 37500.1 (1.6%) | 0.02 |
| **Direct arterial vasodilator** | 3,044 (1.2%) | 12,765 (0.5%) | 0.07 | 1949.7 (0.9%) | 14352.7 (0.6%) | 0.03 |
| **Aldosterone receptor antagonist** | 4,291 (1.7%) | 25,765 (1.1%) | 0.05 | 3476.7 (1.5%) | 27464 (1.2%) | 0.03 |
| **Thiazide diuretic** | 83,718 (34%) | 771,248 (33%) | 0.01 | 72649.3 (32%) | 765806.6 (33%) | 0.02 |
| **Loop diuretic** | 22,236 (9.0%) | 170,144 (7.3%) | 0.06 | 22040.4 (9.79%) | 176139 (7.6%) | 0.08 |
| **Potassium sparing diuretic** | 6,514 (2.6%) | 76,508 (3.3%) | 0.04 | 7499.4 (3.3%) | 74873.6 (3.2%) | 0.008 |
| **Primary care visits** | 3 (1, 5) | 3 (2, 5) | 0.11 | 3 (1, 5) | 3 (2, 5) | 0.01 |
| **Emergency department visits** | 0 (0, 0) | 0 (0, 0) | 0.15 | 0 (0, 0) | 0 (0, 0) | 0.05 |
| **Hospitalizations** | 0 (0, 0) | 0 (0, 0) | 0.17 | 0 (0, 0) | 0 (0, 0) | 0.05 |
| **Healthcare utilization in academic setting** | 158,971 (64%) | 1,548,476 (66%) | 0.05 | 149437.7 (66%) | 1536189.3 (66%) | 0 |
| **Healthcare utilization in rural setting** | 18,153 (7.3%) | 128,154 (5.5%) | 0.07 | 14919.1 (6.6%) | 131392.9 (5.7%) | 0.04 |
| **Number of Veterans go to primary centers** | 22,401 (15,949, 33,006) | 22,662 (17,037, 36,946) | 0.07 | 22477 (16262, 34167) | 22644 (16656, 34167) | 0 |
| **Frailty** |  |  | 0.03 |  |  | 0.04 |
| Frail | 7,528 (3.0%) | 60,055 (2.6%) |  | 7491.2 (3.3%) | 62650.1 (2.7%) |  |
| Non-frail | 240,274 (97%) | 2,269,143 (97%) |  | 218432.1 (97%) | 2253981.9 (97%) |  |

#### eTable 5. Baseline Characteristics of Veterans Before (with imputation) and After Overlap Weighting. Continuous variables are summarized as mean (SD) or median (IQR), and categorical variables as counts (percentages). Absolute standardized mean differences (ASMD) <0.10 indicate acceptable balance.

|  | **Before Overlap Weighting** | | | **After Overlap Weighting** | |
| --- | --- | --- | --- | --- | --- |
| **Variable** | **ARB**  N = 247,802^1^ | **ACEI**  N = 2,329,198^1^ | **ASMD** | **ARB** | **ACEI** |
| **Treatment initiation (years)** | 2010 (2004, 2014) | 2007 (2004, 2011) | 0.32 | 2010 (2004, 2014) | 2010 (2004, 2014) |
| **Age (years)** | 65 (12) | 63 (12) | 0.2 | 65 (12) | 65 (12) |
| **Sex** |  |  | 0.05 |  |  |
| Female | 13616 (5.5%) | 101356 (4.4%) |  | 10004.3 (5.3%) | 10004.3 (5.3%) |
| Male | 234192 (95%) | 2227871 (96%) |  | 179692.3 (95%) | 179692.3 (95%) |
| **Race/Ethnicity** |  |  | 0.06 |  |  |
| Hispanic | 8973 (3.6%) | 107163 (4.6%) |  | 7122.3 (3.8%) | 7122.3 (3.8%) |
| Non-Hispanic Black | 35399 (14%) | 352517 (15%) |  | 26899.4 (14%) | 26899.4 (14%) |
| Non-Hispanic White | 162107 (65%) | 1501083 (64%) |  | 123544.4 (65%) | 123544.4 (65%) |
| Other | 41329 (17%) | 368464 (16%) |  | 32130.6 (17%) | 32130.6 (17%) |
| **County-level income** | 37154 (30702.5, 44648) | 34558 (29007, 41326) | 0.24 | 36726 (30341, 44150) | 36660.5 (30329, 44082) |
| **Veterans integrated service network region** |  |  | 0.17 |  |  |
| Northeast | 49111 (20%) | 462383 (20%) |  | 39370 (21%) | 39370 (21%) |
| Continental | 75525 (30%) | 648561 (28%) |  | 56993.9 (30%) | 56993.9 (30%) |
| Southeast | 72751 (29%) | 848253 (36%) |  | 59129.1 (31%) | 59129.1 (31%) |
| Pacific | 50421 (20%) | 370030 (16%) |  | 34204 (18%) | 34204 (18%) |
| **Supplemental insurance type** |  |  | 0.33 |  |  |
| MEDICAIDE | 363 (0.1%) | 2452 (0.1%) |  | 290.5 (0.2%) | 290.5 (0.2%) |
| MEDICARE | 20485 (8.3%) | 81075 (3.5%) |  | 14618.5 (7.7%) | 14618.5 (7.7%) |
| None | 19582 (7.9%) | 98911 (4.2%) |  | 14651.8 (7.7%) | 14651.8 (7.7%) |
| PRIVATE | 11961 (4.8%) | 43341 (1.9%) |  | 8324 (4.4%) | 8324 (4.4%) |
| Unknown | 195417 (79%) | 2103448 (90%) |  | 151812.1 (80%) | 151812.1 (80%) |
| **Priority group status** |  |  | 0.05 |  |  |
| 1(highest need) | 41806.5 (17%) | 352273.4 (15%) |  | 31931.8 (17%) | 31931.8 (17%) |
| 2 through 9 | 206001.5 (83%) | 1976953.6 (85%) |  | 157765 (83%) | 157765 (83%) |
| **Current smoker** | 28696 (12%) | 444072 (19%) | 0.21 | 23742.1 (13%) | 23742.1 (13%) |
| **Homeless or history of homelessness** | 3690 (1.5%) | 64309 (2.8%) | 0.09 | 3157.8 (1.7%) | 3157.8 (1.7%) |
| **Body mass index, kg/m2** | 31.1 (5.9) | 30.5 (6) | 0.11 | 31 (5.9) | 31 (6.2) |
| **Systolic blood pressure, mm Hg** | 141 (18) | 144 (18) | 0.14 | 142 (18) | 142 (18) |
| **Diastolic blood pressure, mm Hg** | 80 (12) | 82 (12) | 0.2 | 80 (12) | 80 (12) |
| **Heart rate, beats/minute** | 74 (13) | 75 (14) | 0.07 | 74 (13) | 74 (14) |
| **Total cholesterol, mg/dL** | 180 (42) | 186 (43) | 0.14 | 181 (42) | 181 (42) |
| **High-density lipoprotein cholesterol, mg/dL** | 45 (14) | 45 (15) | 0.003 | 45 (14) | 45 (14) |
| **Low-density lipoprotein cholesterol, mg/dL** | 105 (35) | 110 (36) | 0.14 | 105 (35) | 105 (35) |
| **Triglycerides, mg/dL** | 136 (95, 194) | 138 (95, 200) | 0.04 | 137 (95, 196) | 136 (94, 196) |
| **Serum creatinine, mg/dL** | 1.1 (0.9, 1.24) | 1 (0.9, 1.2) | 0.15 | 1.09 (0.9, 1.21) | 1.09 (0.9, 1.2) |
| **Hemoglobin A1c** |  |  | 0.07 |  |  |
| < 5.7% | 27843 (11%) | 248469 (11%) |  | 21483 (11%) | 21483 (11%) |
| 5.7% - < 6.5% | 40990 (17%) | 361935 (16%) |  | 31520.9 (17%) | 31520.9 (17%) |
| 6.5% - < 8.0% | 29480 (12%) | 266695 (11%) |  | 22944.1 (12%) | 22944.1 (12%) |
| 8.0% - < 10.0% | 10984 (4.4%) | 107984 (4.6%) |  | 8695.4 (4.6%) | 8695.4 (4.6%) |
| $\geq$ 10% | 4310 (1.7%) | 58879 (2.5%) |  | 3537.7 (1.9%) | 3537.7 (1.9%) |
| Unknown | 134201 (54%) | 1285265 (55%) |  | 101515.7 (54%) | 101515.7 (54%) |
| **Estimated glomerular filtration rate,**  **ml/min/1.73m^2^** | 77 (62, 92) | 82 (68, 96) | 0.25 | 78 (62, 93) | 77 (63, 92) |
| **Urinary albumin to creatinine ratio** |  |  | 0.03 |  |  |
| < 30 mg/g | 20009 (8.1%) | 185476 (8%) |  | 15498.4 (8.2%) | 15498.4 (8.2%) |
| 30 - < 300 mg/g | 27078 (11%) | 238769 (10%) |  | 21118.4 (11%) | 21118.4 (11%) |
| 300 - < 1000 mg/g | 1404 (0.6%) | 12257 (0.5%) |  | 1098.5 (0.6%) | 1098.5 (0.6%) |
| $\geq$ 1000 mg/g | 177 (<0.1%) | 1076 (<0.1%) |  | 130.8 (<0.1%) | 130.8 (<0.1%) |
| Unknown | 199140 (0.8) | 1891649 (0.81) |  | 151851 (0.8) | 151851 (0.8) |
| **Serum sodium, mEq/L** | 139.3 (2.8) | 139.2 (2.9) | 0.03 | 139.3 (2.83) | 139.3 (2.9) |
| **Serum potassium, mEq/L** | 4.25 (0.45) | 4.24 (0.44) | 0.009 | 4.25 (0.44) | 4.25 (0.44) |
| **Ejection fraction** |  |  | 0.09 |  |  |
| $\leq$ 40% | 8515 (3.4%) | 90202 (3.9%) |  | 7325.7 (3.9%) | 7325.7 (3.9%) |
| 40% - 50% | 5113 (2.1%) | 62835 (2.7%) |  | 4265.8 (2.2%) | 4265.8 (2.2%) |
| > 50% | 22035 (8.9%) | 254581 (11%) |  | 17946.4 (9.5%) | 17946.4 (9.5%) |
| Unknown | 212145 (86%) | 1921609 (82%) |  | 160158.8 (84%) | 160158.8 (84%) |
| **Cardiovascular disease** | 62338 (25%) | 245543 (11%) | 0.39 | 29639.7 (16%) | 29639.7 (16%) |
| **Myocardial infarction** | 597 (0.2%) | 16355 (0.7%) | 0.07 | 537.1 (0.3%) | 537.1 (0.3%) |
| **Peripheral arterial disease** | 3106 (1.3%) | 44638 (1.9%) | 0.05 | 2593.7 (1.4%) | 2593.7 (1.4%) |
| **Any stroke** | 10428 (4.2%) | 106047 (4.6%) | 0.02 | 8241.6 (4.3%) | 8241.6 (4.3%) |
| **Prior coronary revascularization** | 239 (0%) | 6855 (0.3%) | 0.04 | 218 (0.1%) | 218 (0.1%) |
| **Any heart failure** | 54128 (22%) | 135085 (5.8%) | 0.48 | 22422.9 (12%) | 22422.9 (12%) |
| **Alcohol abuse** | 4415 (1.8%) | 89986 (3.9%) | 0.13 | 3758.3 (2%) | 3758.3 (2%) |
| **Angina** | 1034 (0.4%) | 23010 (1%) | 0.07 | 913.9 (0.5%) | 913.9 (0.5%) |
| **Arrhythmia** | 8072 (3.3%) | 82750 (3.6%) | 0.02 | 6524.8 (3.4%) | 6524.8 (3.4%) |
| **Cancer** | 1161 (0.5%) | 17649 (0.8%) | 0.04 | 977.6 (0.5%) | 977.6 (0.5%) |
| **Chronic kidney disease** | 9743 (3.9%) | 65538 (2.8%) | 0.06 | 7355.9 (3.9%) | 7355.9 (3.9%) |
| **Chronic liver disease** | 2019 (0.8%) | 32982 (1.4%) | 0.06 | 1707.4 (0.9%) | 1707.4 (0.9%) |
| **Chronic lung disease** | 12744 (5.1%) | 158047 (6.8%) | 0.07 | 10480 (5.5%) | 10480 (5.5%) |
| **Cirrhosis** | 399 (0.2%) | 6475 (0.3%) | 0.02 | 338.9 (0.2%) | 338.9 (0.2%) |
| **Diabetes** | 37867 (15%) | 398974 (17%) | 0.05 | 30707.7 (16%) | 30707.7 (16%) |
| **Depression** | 19273 (7.8%) | 251127 (11%) | 0.1 | 15644.5 (8.2%) | 15644.5 (8.2%) |
| **Drug/substance use** | 2776 (1.1%) | 56464 (2.4%) | 0.1 | 2381.4 (1.3%) | 2381.4 (1.3%) |
| **Obstructive sleep apnea** | 8577 (3.5%) | 68100 (2.9%) | 0.03 | 6687.6 (3.5%) | 6687.6 (3.5%) |
| **Syncope** | 488 (0.2%) | 8143 (0.3%) | 0.03 | 421.1 (0.2%) | 421.1 (0.2%) |
| **Statin** | 108629 (44%) | 992422 (43%) | 0.02 | 83565.9 (44%) | 83565.9 (44%) |
| **Aspirin** | 27862 (11%) | 327480 (14%) | 0.08 | 22467 (12%) | 22467 (12%) |
| **Sodium-glucose transporter type 2 inhibitor** | 69 (0%) | 173 (0%) | 0.02 | 42 (0%) | 42 (0%) |
| **Antidepressant** | 39377 (16%) | 440385 (19%) | 0.08 | 30955.7 (16%) | 30955.7 (16%) |
| **Number of antihypertensive medications** | 2 (1, 3) | 2 (1, 2) | 0.14 | 2 (1, 3) | 2 (1, 3) |
| **Alpha-blocker** | 20472 (8.3%) | 203550 (8.7%) | 0.02 | 16220.2 (8.58%) | 16220.2 (8.58%) |
| **Beta-blocker** | 70320 (28%) | 651972 (28%) | 0.01 | 54757.6 (29%) | 54757.6 (29%) |
| **Calcium channel blocker** | 63061 (25%) | 459706 (20%) | 0.14 | 47216.1 (25%) | 47216.1 (25%) |
| **Centrally-acting drug** | 5419 (2.2%) | 36319 (1.6%) | 0.05 | 3952.5 (2.1%) | 3952.5 (2.1%) |
| **Direct arterial vasodilator** | 3044 (1.2%) | 12765 (0.5%) | 0.07 | 2091.8 (1.1%) | 2091.8 (1.1%) |
| **Aldosterone receptor antagonist** | 4292 (1.7%) | 25766 (1.1%) | 0.05 | 3185.2 (1.7%) | 3185.2 (1.7%) |
| **Thiazide diuretic** | 83718 (34%) | 771256 (33%) | 0.01 | 62635.1 (33%) | 62635.1 (33%) |
| **Loop diuretic** | 22237 (9%) | 170146 (7.3%) | 0.06 | 17303.2 (9.1%) | 17303.2 (9.1%) |
| **Potassium sparing diuretic** | 6514 (2.6%) | 76509 (3.3%) | 0.04 | 5352.7 (2.8%) | 5352.7 (2.8%) |
| **Primary care visits** | 3 (1, 5) | 3 (2, 5) | 0.11 | 3 (1, 5) | 3 (2, 5) |
| **Emergency department visits** | 0 (0, 0) | 0 (0, 0) | 0.15 | 0 (0, 0) | 0 (0, 0) |
| **Hospitalizations** | 0 (0, 0) | 0 (0, 0) | 0.17 | 0 (0, 0) | 0 (0, 0) |
| **Healthcare utilization in academic setting** | 158975 (64%) | 1548492 (66%) | 0.05 | 122904.5 (65%) | 122904.5 (65%) |
| **Healthcare utilization in rural setting** | 18155 (7.3%) | 128154 (5.5%) | 0.07 | 13532.5 (7.1%) | 13532.5 (7.1%) |
| **Number of Veterans go to primary centers** | 22401 (15949, 33006) | 22662 (17037, 36947) | 0.07 | 22401 (15949, 34167) | 22562 (16388, 33006) |
| **Frailty** |  |  | 0.03 |  |  |
| Frail | 7528 (3%) | 60055 (2.6%) |  | 5855.4 (3.1%) | 6088.1 (3.2%) |
| Non-frail | 240280 (97%) | 2269172 (97%) |  | 183841.4 (97%) | 183608.6 (97%) |

^1^Median (IQR); Mean (SD); n (%)

#### eTable 6. Inverse Probability Weighted Hazard Ratios Comparing ARB vs. ACEI initiators. Cause-specific hazard ratios (HRs) and 95% CIs for dementia identified using NLP algorithms across subgroups.

| **Subgroups** | **Hazard Ratio (95% CI)** | | | | | |
| --- | --- | --- | --- | --- | --- | --- |
|  | 0-0.5 year | 0.5-1 year | 1-2 year | 2-3 year | 3-4 year | 4-5 year |
| **Overall** | 0.88 (0.83, 0.93) | 0.86 (0.8, 0.92) | 0.91 (0.86, 0.95) | 0.97 (0.92, 1.01) | 0.93 (0.89, 0.98) | 0.95 (0.9, 0.99) |
| **Age** | | | | | | |
| <60 | 1.02 (0.88, 1.19) | 0.94 (0.78, 1.12) | 0.94 (0.83, 1.07) | 1.03 (0.91, 1.17) | 0.89 (0.78, 1.01) | 0.93 (0.82, 1.05) |
| >=60 | 0.82 (0.77, 0.87) | 0.81 (0.75, 0.87) | 0.86 (0.82, 0.91) | 0.91 (0.86, 0.96) | 0.9 (0.85, 0.95) | 0.91 (0.86, 0.96) |
| **Sex** | | | | | | |
| Female | 1.11 (0.85, 1.44) | 1.01 (0.74, 1.37) | 1.12 (0.9, 1.4) | 1.1 (0.87, 1.4) | 1.02 (0.81, 1.3) | 0.9 (0.69, 1.16) |
| Male | 0.87 (0.82, 0.92) | 0.85 (0.79, 0.91) | 0.9 (0.85, 0.94) | 0.96 (0.91, 1.01) | 0.93 (0.88, 0.98) | 0.95 (0.9, 1) |
| **Race and ethnicity** | | | | | | |
| Not Black | 0.88 (0.83, 0.94) | 0.85 (0.79, 0.91) | 0.89 (0.85, 0.94) | 0.97 (0.92, 1.02) | 0.92 (0.88, 0.98) | 0.95 (0.9, 1) |
| Black | 0.86 (0.73, 1.01) | 0.88 (0.72, 1.07) | 0.98 (0.85, 1.12) | 0.92 (0.79, 1.06) | 0.97 (0.84, 1.11) | 0.93 (0.81, 1.07) |
| **Systolic blood pressure** | | | | | | |
| <140 | 0.81 (0.74, 0.89) | 0.9 (0.82, 1) | 0.92 (0.86, 0.99) | 0.96 (0.89, 1.04) | 0.91 (0.85, 0.98) | 0.96 (0.89, 1.04) |
| >=140 | 0.94 (0.86, 1.02) | 0.82 (0.74, 0.9) | 0.89 (0.83, 0.95) | 0.97 (0.9, 1.04) | 0.95 (0.88, 1.01) | 0.94 (0.87, 1) |
| **Diabetes** | | | | | | |
| No | 0.88 (0.83, 0.94) | 0.84 (0.78, 0.91) | 0.91 (0.86, 0.96) | 0.98 (0.93, 1.03) | 0.93 (0.88, 0.98) | 0.94 (0.89, 0.99) |
| Yes | 0.87 (0.76, 1) | 0.91 (0.78, 1.06) | 0.89 (0.8, 1) | 0.92 (0.82, 1.03) | 0.95 (0.85, 1.06) | 0.98 (0.88, 1.09) |
| **Number of anti-HTN medications** | | | | | | |
| <2 | 0.86 (0.77, 0.95) | 0.83 (0.73, 0.94) | 0.96 (0.88, 1.05) | 0.95 (0.87, 1.04) | 0.96 (0.88, 1.05) | 1.01 (0.93, 1.1) |
| >=2 | 0.88 (0.82, 0.94) | 0.86 (0.79, 0.93) | 0.87 (0.82, 0.92) | 0.96 (0.9, 1.02) | 0.91 (0.85, 0.96) | 0.9 (0.85, 0.96) |
| **Statin** | | | | | | |
| No | 0.84 (0.77, 0.91) | 0.79 (0.72, 0.87) | 0.87 (0.82, 0.93) | 0.94 (0.88, 1) | 0.93 (0.87, 1) | 0.95 (0.89, 1.02) |
| Yes | 0.93 (0.85, 1.02) | 0.94 (0.85, 1.03) | 0.95 (0.88, 1.02) | 0.99 (0.93, 1.07) | 0.93 (0.86, 1) | 0.94 (0.87, 1.01) |

#### eTable 7. Overlap Weighted Hazard Ratios Comparing ARB vs. ACEI initiators. Cause-specific hazard ratios (HRs) and 95% CIs for dementia identified using NLP algorithms across subgroups.

| **Subgroups** | **Hazard Ratio (95% CI)** | | | | | |
| --- | --- | --- | --- | --- | --- | --- |
|  | 0-0.5 year | 0.5-1 year | 1-2 year | 2-3 year | 3-4 year | 4-5 year |
| **Overall** | 0.82 (0.78, 0.87) | 0.82 (0.77, 0.87) | 0.87 (0.84, 0.91) | 0.92 (0.88, 0.96) | 0.9 (0.86, 0.94) | 0.92 (0.88, 0.96) |
| **Age** | | | | | | |
| <60 | 1.05 (0.92, 1.2) | 0.94 (0.8, 1.11) | 0.94 (0.84, 1.05) | 1.05 (0.94, 1.18) | 0.96 (0.85, 1.08) | 0.97 (0.87, 1.08) |
| >=60 | 0.79 (0.75, 0.84) | 0.8 (0.75, 0.85) | 0.86 (0.82, 0.9) | 0.9 (0.86, 0.94) | 0.89 (0.85, 0.93) | 0.91 (0.87, 0.95) |
| **Sex** | | | | | | |
| Female | 0.97 (0.76, 1.22) | 0.95 (0.73, 1.25) | 1.06 (0.87, 1.29) | 0.89 (0.72, 1.1) | 1.01 (0.82, 1.24) | 0.87 (0.7, 1.08) |
| Male | 0.82 (0.78, 0.86) | 0.81 (0.77, 0.87) | 0.86 (0.83, 0.9) | 0.92 (0.88, 0.96) | 0.9 (0.86, 0.94) | 0.92 (0.88, 0.97) |
| **Race and ethnicity** | | | | | | |
| Not Black | 0.82 (0.77, 0.87) | 0.83 (0.78, 0.88) | 0.87 (0.83, 0.91) | 0.93 (0.89, 0.98) | 0.9 (0.86, 0.94) | 0.93 (0.88, 0.97) |
| Black | 0.87 (0.75, 1) | 0.78 (0.65, 0.92) | 0.9 (0.8, 1.02) | 0.85 (0.75, 0.96) | 0.92 (0.82, 1.04) | 0.89 (0.79, 1.01) |
| **Systolic blood pressure** | | | | | | |
| <140 | 0.79 (0.73, 0.86) | 0.89 (0.82, 0.97) | 0.91 (0.85, 0.97) | 0.95 (0.89, 1.02) | 0.89 (0.83, 0.95) | 0.94 (0.88, 1.01) |
| >=140 | 0.85 (0.79, 0.92) | 0.76 (0.7, 0.83) | 0.84 (0.79, 0.89) | 0.9 (0.84, 0.95) | 0.91 (0.86, 0.97) | 0.9 (0.85, 0.96) |
| **Diabetes** | | | | | | |
| No | 0.82 (0.78, 0.87) | 0.81 (0.76, 0.87) | 0.88 (0.84, 0.93) | 0.93 (0.89, 0.98) | 0.9 (0.85, 0.94) | 0.91 (0.87, 0.96) |
| Yes | 0.84 (0.75, 0.95) | 0.86 (0.76, 0.98) | 0.83 (0.76, 0.92) | 0.88 (0.79, 0.97) | 0.92 (0.84, 1.02) | 0.96 (0.87, 1.06) |
| **Number of anti-HTN medications** | | | | | | |
| <2 | 0.76 (0.7, 0.84) | 0.8 (0.72, 0.89) | 0.91 (0.84, 0.98) | 0.91 (0.85, 0.99) | 0.92 (0.85, 1) | 0.98 (0.9, 1.05) |
| >=2 | 0.85 (0.8, 0.91) | 0.83 (0.77, 0.89) | 0.86 (0.81, 0.9) | 0.92 (0.88, 0.97) | 0.89 (0.85, 0.94) | 0.89 (0.85, 0.94) |
| **Statin** | | | | | | |
| No | 0.79 (0.73, 0.84) | 0.76 (0.7, 0.82) | 0.85 (0.8, 0.9) | 0.89 (0.84, 0.95) | 0.9 (0.85, 0.96) | 0.93 (0.88, 0.99) |
| Yes | 0.88 (0.81, 0.95) | 0.9 (0.82, 0.98) | 0.9 (0.84, 0.96) | 0.95 (0.89, 1.01) | 0.9 (0.85, 0.97) | 0.91 (0.85, 0.97) |

#### eTable 8. Inverse Probability Weighted Risks for Secondary Outcomes among ARB vs. ACEI initiators. Absolute risks, risk ratios, and risk differences for dementia, dementia-free death, and the composite of dementia or death within 5 years. Dementia was identified using diagnosis codes only.

| Outcome | Absolute risk (95% CI) | | Risk Ratio  (95% CI) | Risk Difference  (95% CI) |
| --- | --- | --- | --- | --- |
|  | **ARB initiators** | **ACEI initiators**^1^ |  |  |
| Primary Outcome | | | | |
| Dementia | | | | |
| 0.5-year | 0.001 (0.001, 0.001) | 0.001 (0.001, 0.001) | 0.812 (0.689, 0.956) | 0 (0, 0) |
| 1-year | 0.002 (0.002, 0.002) | 0.002 (0.002, 0.002) | 0.847 (0.754, 0.951) | 0 (-0.001, 0) |
| 2-year | 0.004 (0.004, 0.005) | 0.005 (0.005, 0.005) | 0.864 (0.802, 0.931) | -0.001 (-0.001, 0) |
| 3-year | 0.007 (0.006, 0.007) | 0.008 (0.008, 0.008) | 0.85 (0.798, 0.904) | -0.001 (-0.002, -0.001) |
| 4-year | 0.01 (0.009, 0.01) | 0.011 (0.011, 0.011) | 0.856 (0.812, 0.903) | -0.002 (-0.002, -0.001) |
| 5-year | 0.013 (0.012, 0.013) | 0.014 (0.014, 0.015) | 0.876 (0.835, 0.919) | -0.002 (-0.002, -0.001) |
| Dementia-free death^2^ | | | | |
| 0.5-year | 0.009 (0.009, 0.01) | 0.01 (0.01, 0.01) | 0.898 (0.852, 0.947) | -0.001 (-0.002, -0.001) |
| 1-year | 0.02 (0.02, 0.021) | 0.022 (0.022, 0.022) | 0.921 (0.89, 0.954) | -0.002 (-0.002, -0.001) |
| 2-year | 0.046 (0.045, 0.047) | 0.048 (0.047, 0.048) | 0.96 (0.94, 0.981) | -0.002 (-0.003, -0.001) |
| 3-year | 0.072 (0.071, 0.074) | 0.075 (0.074, 0.075) | 0.968 (0.951, 0.984) | -0.002 (-0.004, -0.001) |
| 4-year | 0.101 (0.099, 0.102) | 0.103 (0.103, 0.104) | 0.976 (0.962, 0.99) | -0.002 (-0.004, -0.001) |
| 5-year | 0.131 (0.13, 0.133) | 0.133 (0.133, 0.134) | 0.986 (0.974, 0.999) | -0.002 (-0.003, 0) |
| Secondary outcomes | | | | |
| Composite of dementia and death | | | | |
| 0.5-year | 0.01 (0.01, 0.011) | 0.011 (0.011, 0.011) | 0.89 (0.849, 0.933) | -0.001 (-0.002, -0.001) |
| 1-year | 0.022 (0.022, 0.023) | 0.024 (0.024, 0.025) | 0.914 (0.885, 0.944) | -0.002 (-0.003, -0.001) |
| 2-year | 0.05 (0.049, 0.051) | 0.053 (0.052, 0.053) | 0.951 (0.931, 0.971) | -0.003 (-0.004, -0.002) |
| 3-year | 0.079 (0.078, 0.08) | 0.083 (0.082, 0.083) | 0.956 (0.941, 0.972) | -0.004 (-0.005, -0.002) |
| 4-year | 0.11 (0.109, 0.112) | 0.114 (0.114, 0.115) | 0.964 (0.951, 0.978) | -0.004 (-0.006, -0.003) |
| 5-year | 0.144 (0.142, 0.146) | 0.148 (0.147, 0.148) | 0.976 (0.964, 0.988) | -0.004 (-0.005, -0.002) |

ACEI: angiotensin-converting enzyme inhibitor; ARB: angiotensin-II receptor blocker; CI: confidence interval; IP: inverse probability

#### eTable 9. Inverse Probability Weighted Risks for Secondary Outcomes among ARB vs. ACEI initiators. Absolute risks, risk ratios, and risk differences for dementia, dementia-free death, and the composite of dementia or death within 5 years. Dementia was identified using diagnosis codes and anti-dementia medication use.

| Outcome | Absolute risk (95% CI) | | Risk Ratio  (95% CI) | Risk Difference  (95% CI) |
| --- | --- | --- | --- | --- |
|  | **ARB initiators** | **ACEI initiators**^1^ |  |  |
| Primary Outcome | | | | |
| Dementia | | | | |
| 0.5-year | 0.003 (0.003, 0.003) | 0.003 (0.003, 0.003) | 0.947 (0.868, 1.033) | 0 (0, 0) |
| 1-year | 0.005 (0.005, 0.006) | 0.006 (0.006, 0.006) | 0.916 (0.859, 0.976) | 0 (-0.001, 0) |
| 2-year | 0.011 (0.01, 0.011) | 0.011 (0.011, 0.011) | 0.94 (0.895, 0.987) | -0.001 (-0.001, 0) |
| 3-year | 0.016 (0.015, 0.016) | 0.017 (0.017, 0.017) | 0.933 (0.895, 0.971) | -0.001 (-0.002, 0) |
| 4-year | 0.021 (0.021, 0.022) | 0.023 (0.022, 0.023) | 0.946 (0.912, 0.981) | -0.001 (-0.002, 0) |
| 5-year | 0.027 (0.026, 0.028) | 0.028 (0.028, 0.029) | 0.953 (0.923, 0.985) | -0.001 (-0.002, 0) |
| Dementia-free death^2^ | | | | |
| 0.5-year | 0.009 (0.009, 0.01) | 0.01 (0.01, 0.01) | 0.898 (0.852, 0.947) | -0.001 (-0.002, -0.001) |
| 1-year | 0.02 (0.019, 0.021) | 0.022 (0.022, 0.022) | 0.92 (0.889, 0.953) | -0.002 (-0.002, -0.001) |
| 2-year | 0.045 (0.044, 0.046) | 0.047 (0.047, 0.047) | 0.96 (0.939, 0.981) | -0.002 (-0.003, -0.001) |
| 3-year | 0.071 (0.07, 0.072) | 0.074 (0.073, 0.074) | 0.967 (0.951, 0.984) | -0.002 (-0.004, -0.001) |
| 4-year | 0.099 (0.097, 0.1) | 0.101 (0.101, 0.102) | 0.975 (0.961, 0.99) | -0.003 (-0.004, -0.001) |
| 5-year | 0.128 (0.127, 0.13) | 0.13 (0.13, 0.131) | 0.986 (0.973, 0.999) | -0.002 (-0.004, 0) |
| Secondary outcomes | | | | |
| Composite of dementia and death | | | | |
| 0.5-year | 0.012 (0.011, 0.012) | 0.013 (0.013, 0.013) | 0.909 (0.871, 0.949) | -0.001 (-0.002, -0.001) |
| 1-year | 0.025 (0.025, 0.026) | 0.028 (0.027, 0.028) | 0.919 (0.893, 0.947) | -0.002 (-0.003, -0.001) |
| 2-year | 0.056 (0.055, 0.057) | 0.058 (0.058, 0.059) | 0.956 (0.937, 0.976) | -0.003 (-0.004, -0.001) |
| 3-year | 0.087 (0.086, 0.088) | 0.091 (0.09, 0.091) | 0.961 (0.945, 0.976) | -0.004 (-0.005, -0.002) |
| 4-year | 0.12 (0.119, 0.122) | 0.124 (0.124, 0.124) | 0.97 (0.957, 0.983) | -0.004 (-0.005, -0.002) |
| 5-year | 0.155 (0.154, 0.157) | 0.158 (0.158, 0.159) | 0.98 (0.968, 0.992) | -0.003 (-0.005, -0.001) |

ACEI: angiotensin-converting enzyme inhibitor; ARB: angiotensin-II receptor blocker; CI: confidence interval; IP: inverse probability

#### eTable 10. Overlap Weighted Risks for Primary and Secondary Outcomes among ARB vs. ACEI initiators. Absolute risks, risk ratios, and risk differences for dementia, dementia-free death, and the composite of dementia or death within 5 years. Dementia was identified using a validated natural language processing (NLP) algorithm.

| Outcome | Absolute risk (95% CI) | | Risk Ratio  (95% CI) | Risk Difference  (95% CI) |
| --- | --- | --- | --- | --- |
|  | **ARB initiators** | **ACEI initiators**^1^ |  |  |
| Primary Outcome | | | | |
| Dementia | | | | |
| 0.5-year | 0.007 (0.007, 0.008) | 0.009 (0.009, 0.009) | 0.826 (0.787, 0.867) | -0.002 (-0.002, -0.001) |
| 1-year | 0.013 (0.013, 0.014) | 0.016 (0.016, 0.016) | 0.825 (0.795, 0.856) | -0.003 (-0.003, -0.002) |
| 2-year | 0.024 (0.023, 0.025) | 0.028 (0.028, 0.029) | 0.849 (0.825, 0.874) | -0.004 (-0.005, -0.004) |
| 3-year | 0.035 (0.034, 0.036) | 0.040 (0.039, 0.040) | 0.874 (0.854, 0.894) | -0.005 (-0.006, -0.004) |
| 4-year | 0.045 (0.044, 0.046) | 0.051 (0.051, 0.052) | 0.884 (0.865, 0.902) | -0.006 (-0.007, -0.005) |
| 5-year | 0.056 (0.055, 0.057) | 0.062 (0.062, 0.063) | 0.894 (0.877, 0.910) | -0.007 (-0.008, -0.006) |
| Dementia-free death^2^ | | | | |
| 0.5-year | 0.009 (0.008, 0.009) | 0.011 (0.010, 0.011) | 0.836 (0.797, 0.878) | -0.002 (-0.002, -0.001) |
| 1-year | 0.019 (0.019, 0.020) | 0.023 (0.022, 0.023) | 0.845 (0.819, 0.872) | -0.004 (-0.004, -0.003) |
| 2-year | 0.043 (0.043, 0.044) | 0.049 (0.048, 0.049) | 0.888 (0.870, 0.906) | -0.005 (-0.006, -0.005) |
| 3-year | 0.068 (0.067, 0.069) | 0.076 (0.076, 0.077) | 0.894 (0.880, 0.908) | -0.008 (-0.009, -0.007) |
| 4-year | 0.095 (0.094, 0.096) | 0.104 (0.103, 0.105) | 0.912 (0.900, 0.925) | -0.009 (-0.010, -0.008) |
| 5-year | 0.123 (0.122, 0.125) | 0.133 (0.132, 0.134) | 0.926 (0.915, 0.936) | -0.010 (-0.011, -0.008) |
| Secondary outcomes | | | | |
| Composite of dementia and death | | | | |
| 0.5-year | 0.016 (0.016, 0.017) | 0.020 (0.019, 0.020) | 0.832 (0.805, 0.859) | -0.003 (-0.004, -0.003) |
| 1-year | 0.032 (0.032, 0.033) | 0.039 (0.038, 0.039) | 0.837 (0.818, 0.857) | -0.006 (-0.007, -0.006) |
| 2-year | 0.067 (0.066, 0.068) | 0.077 (0.076, 0.078) | 0.874 (0.859, 0.888) | -0.010 (-0.011, -0.009) |
| 3-year | 0.103 (0.102, 0.104) | 0.116 (0.115, 0.117) | 0.887 (0.876, 0.898) | -0.013 (-0.014, -0.012) |
| 4-year | 0.140 (0.139, 0.142) | 0.155 (0.154, 0.156) | 0.903 (0.893, 0.913) | -0.015 (-0.017, -0.014) |
| 5-year | 0.179 (0.177, 0.181) | 0.195 (0.194, 0.196) | 0.916 (0.907, 0.924) | -0.017 (-0.018, -0.015) |
| All-cause death | | | | |
| 0.5-year | 0.009 (0.009, 0.009) | 0.011 (0.011, 0.011) | 0.827 (0.789, 0.867) | -0.002 (-0.002, -0.001) |
| 1-year | 0.020 (0.020, 0.021) | 0.024 (0.024, 0.024) | 0.836 (0.811, 0.862) | -0.004 (-0.005, -0.003) |
| 2-year | 0.046 (0.046, 0.047) | 0.053 (0.052, 0.053) | 0.877 (0.860, 0.894) | -0.007 (-0.007, -0.006) |
| 3-year | 0.074 (0.073, 0.075) | 0.084 (0.083, 0.085) | 0.883 (0.869, 0.896) | -0.010 (-0.011, -0.009) |
| 4-year | 0.105 (0.104, 0.106) | 0.117 (0.116, 0.117) | 0.899 (0.888, 0.910) | -0.012 (-0.013, -0.010) |
| 5-year | 0.137 (0.136, 0.139) | 0.151 (0.150, 0.152) | 0.909 (0.900, 0.919) | -0.014 (-0.015, -0.012) |

ACEI: angiotensin-converting enzyme inhibitor; ARB: angiotensin-II receptor blocker; CI: confidence interval; IP: inverse probability

#### eTable 11. Overlap Weighted Risks for Secondary Outcomes among ARB vs. ACEI initiators. Absolute risks, risk ratios, and risk differences for dementia, dementia-free death, and the composite of dementia or death within 5 years. Dementia was identified using diagnosis codes only.

| Outcome | Absolute risk (95% CI) | | Risk Ratio  (95% CI) | Risk Difference  (95% CI) |
| --- | --- | --- | --- | --- |
|  | **ARB initiators** | **ACEI initiators**^1^ |  |  |
| Primary Outcome | | | | |
| Dementia | | | | |
| 0.5-year | 0.001 (0.001, 0.001) | 0.001 (0.001, 0.001) | 0.752 (0.648, 0.872) | 0 (0, 0) |
| 1-year | 0.002 (0.002, 0.002) | 0.003 (0.002, 0.003) | 0.781 (0.704, 0.868) | -0.001 (-0.001, 0) |
| 2-year | 0.004 (0.004, 0.005) | 0.006 (0.005, 0.006) | 0.8 (0.748, 0.855) | -0.001 (-0.001, -0.001) |
| 3-year | 0.007 (0.007, 0.007) | 0.009 (0.009, 0.009) | 0.797 (0.754, 0.844) | -0.002 (-0.002, -0.001) |
| 4-year | 0.01 (0.009, 0.01) | 0.012 (0.012, 0.012) | 0.808 (0.77, 0.848) | -0.002 (-0.003, -0.002) |
| 5-year | 0.013 (0.012, 0.013) | 0.016 (0.015, 0.016) | 0.824 (0.789, 0.86) | -0.003 (-0.003, -0.002) |
| Dementia-free death^2^ | | | | |
| 0.5-year | 0.009 (0.009, 0.009) | 0.011 (0.011, 0.011) | 0.828 (0.79, 0.868) | -0.002 (-0.002, -0.001) |
| 1-year | 0.02 (0.019, 0.02) | 0.024 (0.023, 0.024) | 0.837 (0.812, 0.863) | -0.004 (-0.005, -0.003) |
| 2-year | 0.046 (0.045, 0.046) | 0.052 (0.051, 0.052) | 0.879 (0.862, 0.897) | -0.006 (-0.007, -0.005) |
| 3-year | 0.072 (0.071, 0.074) | 0.082 (0.081, 0.082) | 0.886 (0.873, 0.9) | -0.009 (-0.01, -0.008) |
| 4-year | 0.102 (0.101, 0.103) | 0.113 (0.112, 0.114) | 0.903 (0.892, 0.915) | -0.011 (-0.012, -0.01) |
| 5-year | 0.133 (0.132, 0.134) | 0.145 (0.145, 0.146) | 0.915 (0.905, 0.925) | -0.012 (-0.014, -0.011) |
| Secondary outcomes | | | | |
| Composite of dementia and death | | | | |
| 0.5-year | 0.01 (0.01, 0.01) | 0.012 (0.012, 0.012) | 0.82 (0.787, 0.855) | -0.002 (-0.003, -0.002) |
| 1-year | 0.022 (0.021, 0.023) | 0.026 (0.026, 0.027) | 0.832 (0.809, 0.855) | -0.004 (-0.005, -0.004) |
| 2-year | 0.05 (0.049, 0.051) | 0.057 (0.057, 0.058) | 0.872 (0.856, 0.888) | -0.007 (-0.008, -0.006) |
| 3-year | 0.079 (0.078, 0.081) | 0.091 (0.09, 0.091) | 0.878 (0.865, 0.891) | -0.011 (-0.012, -0.01) |
| 4-year | 0.112 (0.111, 0.113) | 0.125 (0.124, 0.126) | 0.894 (0.883, 0.905) | -0.013 (-0.015, -0.012) |
| 5-year | 0.146 (0.145, 0.147) | 0.161 (0.16, 0.162) | 0.906 (0.897, 0.916) | -0.015 (-0.017, -0.014) |

ACEI: angiotensin-converting enzyme inhibitor; ARB: angiotensin-II receptor blocker; CI: confidence interval

#### eTable 12. Overlap Weighted Risks for Secondary Outcomes among ARB vs. ACEI initiators. Absolute risks, risk ratios, and risk differences for dementia, dementia-free death, and the composite of dementia or death within 5 years. Dementia was identified using diagnosis codes and anti-dementia medication use.

| Outcome | Absolute risk (95% CI) | | Risk Ratio  (95% CI) | Risk Difference  (95% CI) |
| --- | --- | --- | --- | --- |
|  | **ARB initiators** | **ACEI initiators**^1^ |  |  |
| Primary Outcome | | | | |
| Dementia | | | | |
| 0.5-year | 0.003 (0.003, 0.003) | 0.003 (0.003, 0.003) | 0.862 (0.795, 0.935) | 0 (-0.001, 0) |
| 1-year | 0.005 (0.005, 0.006) | 0.006 (0.006, 0.007) | 0.849 (0.802, 0.899) | -0.001 (-0.001, -0.001) |
| 2-year | 0.011 (0.01, 0.011) | 0.012 (0.012, 0.013) | 0.874 (0.838, 0.911) | -0.002 (-0.002, -0.001) |
| 3-year | 0.016 (0.016, 0.017) | 0.018 (0.018, 0.019) | 0.878 (0.847, 0.909) | -0.002 (-0.003, -0.002) |
| 4-year | 0.022 (0.021, 0.023) | 0.025 (0.024, 0.025) | 0.889 (0.86, 0.918) | -0.003 (-0.003, -0.002) |
| 5-year | 0.028 (0.027, 0.028) | 0.031 (0.031, 0.031) | 0.897 (0.872, 0.923) | -0.003 (-0.004, -0.002) |
| Dementia-free death^2^ | | | | |
| 0.5-year | 0.009 (0.009, 0.009) | 0.011 (0.011, 0.011) | 0.828 (0.79, 0.868) | -0.002 (-0.002, -0.001) |
| 1-year | 0.02 (0.019, 0.02) | 0.024 (0.023, 0.024) | 0.836 (0.811, 0.862) | -0.004 (-0.005, -0.003) |
| 2-year | 0.045 (0.044, 0.046) | 0.051 (0.051, 0.052) | 0.879 (0.861, 0.897) | -0.006 (-0.007, -0.005) |
| 3-year | 0.071 (0.07, 0.072) | 0.081 (0.08, 0.081) | 0.886 (0.872, 0.9) | -0.009 (-0.01, -0.008) |
| 4-year | 0.1 (0.099, 0.101) | 0.111 (0.11, 0.111) | 0.903 (0.891, 0.915) | -0.011 (-0.012, -0.009) |
| 5-year | 0.13 (0.129, 0.131) | 0.142 (0.141, 0.143) | 0.915 (0.904, 0.925) | -0.012 (-0.014, -0.011) |
| Secondary outcomes | | | | |
| Composite of dementia and death | | | | |
| 0.5-year | 0.012 (0.011, 0.012) | 0.014 (0.014, 0.014) | 0.836 (0.804, 0.869) | -0.002 (-0.003, -0.002) |
| 1-year | 0.025 (0.025, 0.026) | 0.03 (0.03, 0.03) | 0.839 (0.819, 0.86) | -0.005 (-0.005, -0.004) |
| 2-year | 0.056 (0.055, 0.057) | 0.064 (0.063, 0.064) | 0.878 (0.863, 0.894) | -0.008 (-0.009, -0.007) |
| 3-year | 0.087 (0.086, 0.089) | 0.099 (0.098, 0.1) | 0.884 (0.872, 0.897) | -0.011 (-0.013, -0.01) |
| 4-year | 0.122 (0.12, 0.123) | 0.135 (0.134, 0.136) | 0.9 (0.89, 0.911) | -0.013 (-0.015, -0.012) |
| 5-year | 0.158 (0.156, 0.159) | 0.173 (0.172, 0.174) | 0.911 (0.902, 0.921) | -0.015 (-0.017, -0.014) |

ACEI: angiotensin-converting enzyme inhibitor; ARB: angiotensin-II receptor blocker; CI: confidence interval

#### eTable 13. Missing Values in Baseline Characteristics. Proportion of missing data for each baseline covariate among all Veterans and by treatment group.

| **Variable** | **Overall**  N = 2,577,000^1^ | **ARB**  N = 247,802^1^ | **ACEI**  N = 2,329,198^1^ |
| --- | --- | --- | --- |
| **County-level income** | 221,476 (8.6%) | 19,623 (7.9%) | 201,853 (8.7%) |
| **Priority group status** | 240,049 (9.3%) | 26,543 (11%) | 213,506 (9.2%) |
| **Body mass index, kg/m2** | 431,813 (17%) | 40,296 (16%) | 391,517 (17%) |
| **Systolic blood pressure, mm Hg** | 257,013 (10.0%) | 24,294 (9.8%) | 232,719 (10.0%) |
| **Diastolic blood pressure, mm Hg** | 257,013 (10.0%) | 24,294 (9.8%) | 232,719 (10.0%) |
| **Heart rate, beats/minute** | 48,666 (1.9%) | 5,671 (2.3%) | 42,995 (1.8%) |
| **Total cholesterol, mg/dL** | 631,415 (25%) | 72,171 (29%) | 559,244 (24%) |
| **High-density lipoprotein cholesterol, mg/dL** | 698,739 (27%) | 77,019 (31%) | 621,720 (27%) |
| **Low-density lipoprotein cholesterol, mg/dL** | 758,085 (29%) | 83,205 (34%) | 674,880 (29%) |
| **Triglycerides, mg/dL** | 714,653 (28%) | 80,308 (32%) | 634,345 (27%) |
| **Serum creatinine, mg/dL** | 446,700 (17%) | 56,438 (23%) | 390,262 (17%) |
| **Estimated glomerular filtration rate, ml/min/1.73m^2** | 446,525 (17%) | 56,421 (23%) | 390,104 (17%) |
| **Serum sodium, mEq/L** | 509,577 (20%) | 62,170 (25%) | 447,407 (19%) |
| **Serum potassium, mEq/L** | 428,959 (17%) | 55,578 (22%) | 373,381 (16%) |
